# Extracorporeal shock wave therapy for erectile dysfunction: rethinking study design, implementation, and analysis

**DOI:** 10.1101/2024.12.10.24318762

**Authors:** Janak Desai, Eric Huyghe, Gayle D. Maffulli, Carmen Nussbaum-Krammer, Jessica Tittelmeier, Christoph Schmitz

**Affiliations:** Department of Urology, Samved Hospital, Ahmedabad, India; Department of Reproductive Medicine, Paule-de-Viguier Hospital, Toulouse University Hospital, Toulouse, France; Department of Urology, Andrology and Renal Transplantation, Rangueil Hospital, Toulouse University Hospital, Toulouse, France; UMR DEFE Inserm 1203, University of Toulouse 3, Toulouse, France; SportsMed UK, London, UK; Department of Anatomy II, Faculty of Medicine, LMU Munich, 80336 Munich, Germany

**Author notes:** **Correspondence** Christoph Schmitz, Chair of Anatomy II, Institute of Anatomy, Faculty of Medicine, Ludwig-Maximilians University, Pettenkoferstr. 11, 80336 Munich, Germany.

**Keywords:** acoustic pressure fields, clinical trials, extracorporeal shock wave therapy, ESWT, erectile dysfunction, estimand, intercurrent events, missing data imputation

## Abstract

**Introduction:** Extracorporeal shock wave therapy (ESWT) for erectile dysfunction (ED) presents a challenging paradox. While numerous clinical studies, systematic reviews, and meta-analyses have been published, indicating a substantial body of evidence supporting the efficacy and safety of ESWT, significant questions remain. Notably, the American Urological Association (AUA) continues to classify ESWT for ED as investigational (Evidence Level: Grade C), suggesting that the true therapeutic effect o f ESWT may differ considerably from current estimates. This review aims to critically assess the evidence and propose strategies to address this unresolved discrepancy.

**Data sources:** We systematically searched two electronic databases (PubMed and Ovid/Embase) and published systematic reviews on ESWT for ED and compiled a systematic literature review and meta-analysis based on 87 relevant studies.

**Areas of agreement:** There is clear evidence that ESWT for ED is effective and can therefore be a valuable treatment modality in the management of ED.

**Areas of controversy:** Current assessments of ESWT for ED as investigational by, e.g., the AUA may not stem from a lack of clinical studies, insufficient related basic science, or an inadequate number of systematic reviews and meta-analyses. Instead, the deficits lie in the area of the scientific quality of the clinical studies published to date.

**Growing points:** We hypothesize that this unfortunate situation will only change if the following aspects will be rigorously considered in future clinical studies on ESWT for ED: adequate characterization and reporting of extracorporeal shock waves, appropriate handling of missing data and intercurrent events, and comprehensive classification of ESWT in the overall context of the available treatment options for ED.

**Areas for developing research:** We are convinced that the consistent implementation of these aspects will significantly contribute to establishing ESWT as the first truly regenerative therapy in the management of ED. This overall aim justifies the corresponding efforts, for the benefit of our patients.

## INTRODUCTION

Erectile dysfunction (ED) is a widespread condition that mainly affects men over 40 years of age and is increasing in prevalence worldwide.^1-3^ It is characterized by the constant or repeated inability to achieve and maintain an erection sufficient for satisfactory sexual performance. The pathophysiology of ED is complex and includes various organic, psychogenic, and mixed factors.^3^ The etiology of ED is often associated with comorbidities, including cardiovascular disease and diabetes.^2-4^ Furthermore, neurological disorders and conditions after radical prostatectomy may play important roles in the development of ED.^2,3^ Untreated ED may lead to low self-esteem, strained interpersonal relationships, anxiety and depression.^2,3^

The treatment modalities for ED recommended in guidelines of, e.g., the European Urological Association (EUA) and the American Urological Association (AUA) include Phosphodiesterase-5 inhibitors (PDE5i), inflatable penile prostheses, vacuum erection devices, intracavernosal injections of alprostadil or other drugs, and intraurethral administration of alprostadil.^5-7^ However, none of these treatment modalities can be considered a curative and regenerative therapy of ED.

Over the last 15 years extracorporeal shock wave therapy (ESWT) has emerged as an attractive, non-invasive treatment modality in the management of ED.^3,7,8^ Originally developed for cracking kidney stones (extracorporeal shock wave lithotripsy),^9^ ESWT has been successfully used for over 30 years to manage a variety of pathologies of the musculoskeletal system.^10-12^ Numerous molecular and cellular mechanisms of action of extracorporeal shock waves (ESWs) on bone and cartilage tissue, connective tissue as well as muscle and nerve tissue have been identified,^13^ making it attractive to use ESWT also in pathologies other than those of the musculoskeletal system. The ability of ESWs to induce functional angiogenesis is of particular relevance in this regard.^13-15^

However, current evaluations of ESWT for ED in the literature and among medical society guidelines come to different conclusions. Sixteen systematic reviews and meta-analyses of clinical studies on ESWT published between 2017 and 2024 (summarized in Table 1) largely indicate that ESWT is an effective intervention for ED.^16-31^ Furthermore, the European Association of Urologists recommends (albeit with weak rating) the use of ESWT with/without PDE5i in patients (i) with mild vasculogenic ED, (ii) as an alternative therapy in well-informed patients who do not wish to have or are not suitable for oral vasoactive therapy, and/or (iii) who are vasculogenic ED patients who are poor responders to PDE5i.^32^ ESWT for ED is also recommended by the European Society for Sexual Medicine.^5^

**Table 1.** Conclusions regarding the efficacy of extracorporeal shock wave therapy for erectile dysfunction in systematic reviews and me ta-analyses published between 2017 and 2024.

| R | NFA | Y-P | Conclusion |
| --- | --- | --- | --- |
| 16 | Hinojosa-Gonzalez | 2024 | Our network meta-analysis suggests that low-intensity extracorporeal shockwave therapy is an effective intervention for erectile dysfunction, as measured by increases in the IIEF-EF. |
| 17 | Vieiralves | 2023 | The literature presents little scientific evidence but suggests good results with the use of LI-EST for ED. |
| 18 | Mason | 2023 | The examined studies present encouraging results for the use of LI-ESWT to treat diabetic men with ED. |
| 19 | Liu | 2022 | Compared with placebo treatment, LI-ESWT alleviates ED symptoms in patients, particularly those who have mild or moderate ED. |
| 20 | Rho | 2022 | The results of this analysis indicate that LI-ESWT showed a statistically significant effect on early recovery in penile rehabilitation of ED following RP. |
| 21 | Yao | 2022 | The results of this meta-analysis suggest that treatment plans with an energy density of 0.09 mJ/mm<sup>2</sup> and pulses number of 1,500 to 2,000 are more beneficial to IIEF in ED patients. In addition, IIEF improvement was more pronounced in patients with moderate ED after extracorporeal shockwave therapy. |
| 22 | Canguven | 2021 | The present review found that LI-ESWT has a role in ED treatment in laboratory studies, but its role in human clinical trials is still controversial. |
| 23 | Kalka | 2021 | Evidence exists that LI-ESWT generated with an electrohydraulic unit is effective. |
| 24 | Ochoa | 2021 | SW may have a theoretical impact on the vascular etiology of organic ED. |
| 25 | Dong | 2019 | In meta-analysis of seven RCTs with men who received LI-ESWT for ED, there was evidence that the IIEF-EF and EHS experienced improvements following LI-ESWT. |
| 26 | Sokolakis | 2019 | The present meta-analysis provided results showing that LI-ESWT significantly improves erectile function in patients with vasculogenic ED. |
| 27 | Campbell | 2019 | This therapeutic strategy appears to be well tolerated with short-term benefits. |
| 28 | Man | 2018 | These studies suggest that LI-ESWT could significantly improve the IIEF and EHS of patients with ED. |
| 29 | Angulo | 2017 | According to the literature, treatment with LI-ESWT for erectile dysfunction is effective, both in the short and medium term. LI-ESWT has been described as more effective than placebo in the short term. |
| 30 | Lu | 2017 | The number of studies of LI-ESWT for ED have increased dramatically in recent years. Most of these studies presented encouraging results, regardless of variation in LI-ESWT setup parameters or treatment protocols. These studies suggest that LI-ESWT could significantly improve the IIEF and EHS of ED patients. |
| 31 | Clavijo | 2017 | In this meta-analysis of seven randomized controlled trials, treatment of ED with LI-ESWT resulted in a significant increase in IIEF-EF scores. |

Abbreviations: R, reference; N-FA, name of the first author; Y-P, year of publication; IIEF, International Index of Erectile Function; IIEF-EF, IIEF - Erectile Domain; ED, erectile dysfunction; RP, radical prostatectomy; SW, shock waves; RCTs, randomized controlled trials; EHS, Erection Hardness Score.

In contrast, the most recent guidelines of the AUA on ED designate ESWT for ED as investigational (Conditional Recommendation; Evidence Level: Grade C).^6^ Evidence Level Grade C is defined by the AUA as low (“our confidence in the effect estimate is limited / the true effect may be substantially different from the estimate of the effect”) or very low (“we have very little confidence in the effect estimate / the true effect is likely to be substantially different from the estimate of effect”).^33^

A comprehensive assessment of the aforementioned systematic reviews and meta-analyses^16-31^ indicate that many studies were excluded from further analysis in these reviews, leaving several critical aspects insufficiently addressed. Specifically, three primary aspects were not, or only barely, critically addressed: (i) the characterization of the different types of ESWs applied in ESWT for ED (focused / unfocused / linear / radial ESWs) where energy density values were frequently mentioned without clarification of whether these values represent positive or total energy density, and where in the three-dimensional (3D) acoustic pressure field of an ESW this energy density is reached, (ii) strategies for managing missing data and intercurrent events in clinical studies on ESWT for ED, and (iii) the integration of ESWT within the broader spectrum of available treatments for ED, particularly with respect to optimizing therapy through targeted combination of several treatment modalities.

Based on these observations we performed a systematic review of clinical studies on ESWT for ED that were published until September 27^th^, 2024, with a particular focus on the aforementioned topics and without excluding those studies that were not suitable for a meta-analysis. The conclusion of our systematic assessment of 87 studies extends beyond the conditional recommendation (investigational) of the AUA: we recommend a fundamental re-evaluation in the design, implementation and analysis of clinical studies on ESWT for ED.

## METHODS

A systematic search was conducted on PubMed and Ovid/Embase using the terms “erectile dysfunction shock wave” and “erectile dysfunction shockwave” from the days of inception of these databases until September 27, 2024, according to the 2020 PRISMA (Preferred Reporting Items for Systematic Reviews and Meta-Analyses)^34^ guidelines. The strategy of the assessment of the identified reports is summarized in Figure 1. For PubMed these searches retrieved 270 and 199 records, respectively. In Ovid/Embase, both searches led to the proposed subject headings “erectile dysfunction” and “shock wave therapy”. Combining these subject headings in a single search with Boolean operator AND yielded 110 records. Thus, the total number of records identified from databases was 579, of which 220 were duplicates that were removed before screening. Furthermore, the 16 meta-analyses listed in Table 1 were searched for studies that were not found in PubMed and Ovid/Embase. After removal of abstracts of presentations at scientific conferences, one additional record remained. Automation tools for marking records as ineligible were not used.

**Figure 1.**
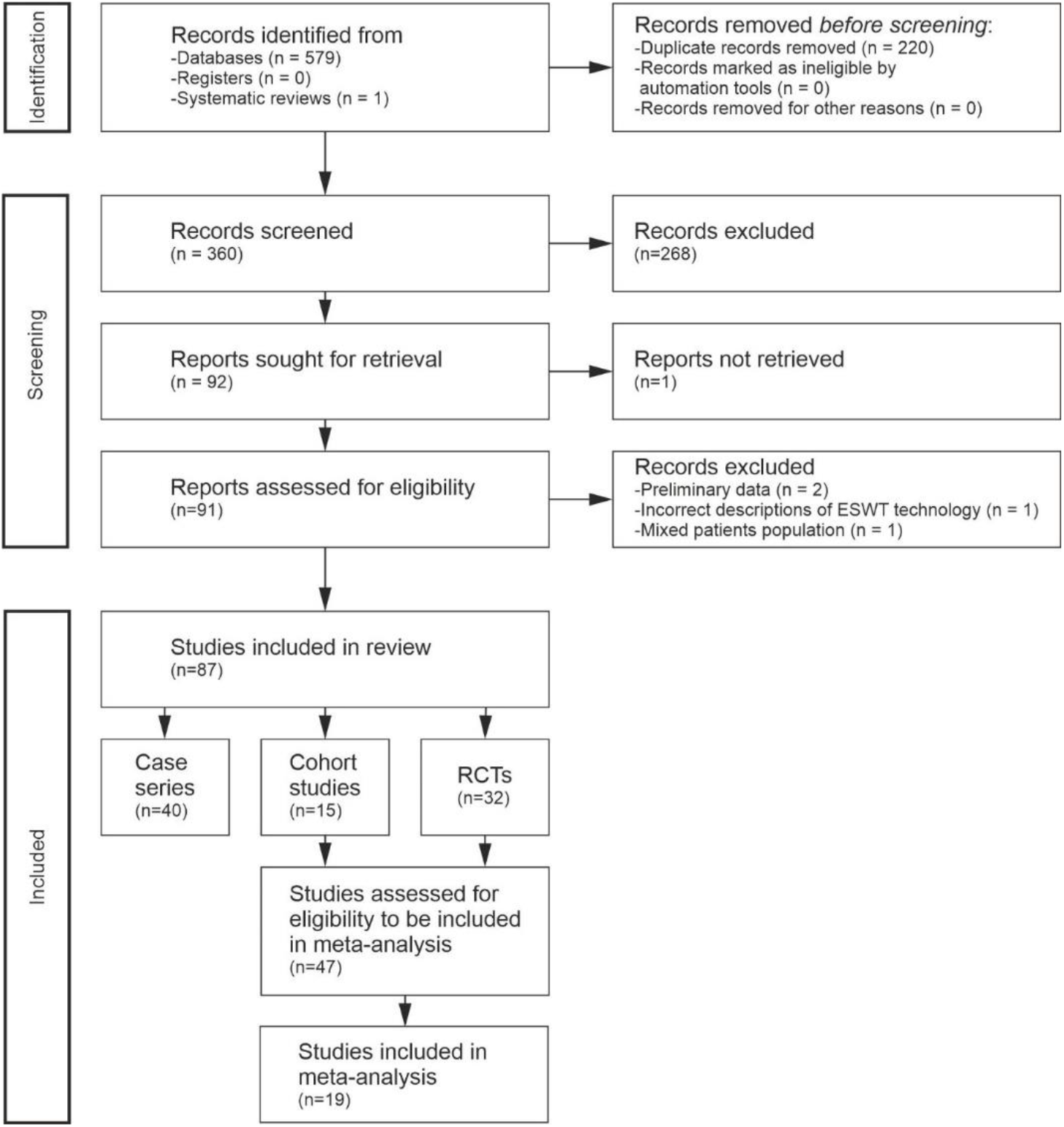
Systematic review flow chart of the literature search regarding extracorporeal shock wave therapy for erectile dysfunction, p erformed according to the PRISMA guidelines^34^ on September 27, 2024.

The resulting 360 records were screened, and studies on other pathologies than ED (e.g., Peyronie’s disease), reviews, commentaries, editorials, letters to the editor and studies on animal models were excluded (together 268 records). From the 92 reports sought for retrieval (references in Supplementary Data), all but one were downloaded from the E-media library of LMU Munich (Munich, Germany) or obtained through other sources, and were assessed for eligibility. Translation from languages other than English was performed using Google Translate. One case series^35^ (n=32 patients; no control group) in Chinese language was not retrieved.

Four of the 91 reports assessed for eligibility were excluded from further analysis. (i) Two publications^36,37^ were preliminary reports (named clinical trial updates by the authors) of a randomized controlled trial (RCT)^38^ after enrollment of approximately 25%^36^ or 50%^37^ of the patients into the RCT. (ii) One publication^39^ was a national, multi-institutional, retrospective progress report about the use of four different ESWT devices in treatment of ED. However, the partially incorrect descriptions of the ESWT devices in this report^39^ raised doubts about the scientific integrity of the entire publication. Specifically, the BTL-6000 SWT device (BTL, Prague, Czech Republic) was described as electropneumatic generator with focused-wave morphology but actually generates radial ESWs (rESWs), whereas the 250 device (Shenzen Huikang Medical Apparatus Co., Shenzen, China) was described as electromagnetic generator with radial-wave morphology but actually generates focused ESWs (fESWs). (iii) In one study^40^ only approximately two thirds of the patients suffered from ED, without separate reporting of the results of the ED patients.

The remaining 87 reports were included in the systematic review. 40 of them were reports of case series without control groups, 15 were reports of cohort studies with one or more, non-randomized control groups and 32 were reports of RCTs. Table 2 summarizes the variables that were extracted from the 87 identified reports. For case series, 38 variables were extracted, and for cohort studies and RCTs with one / two / three control groups 45 / 50 / 55 variables.

**Table 2.**
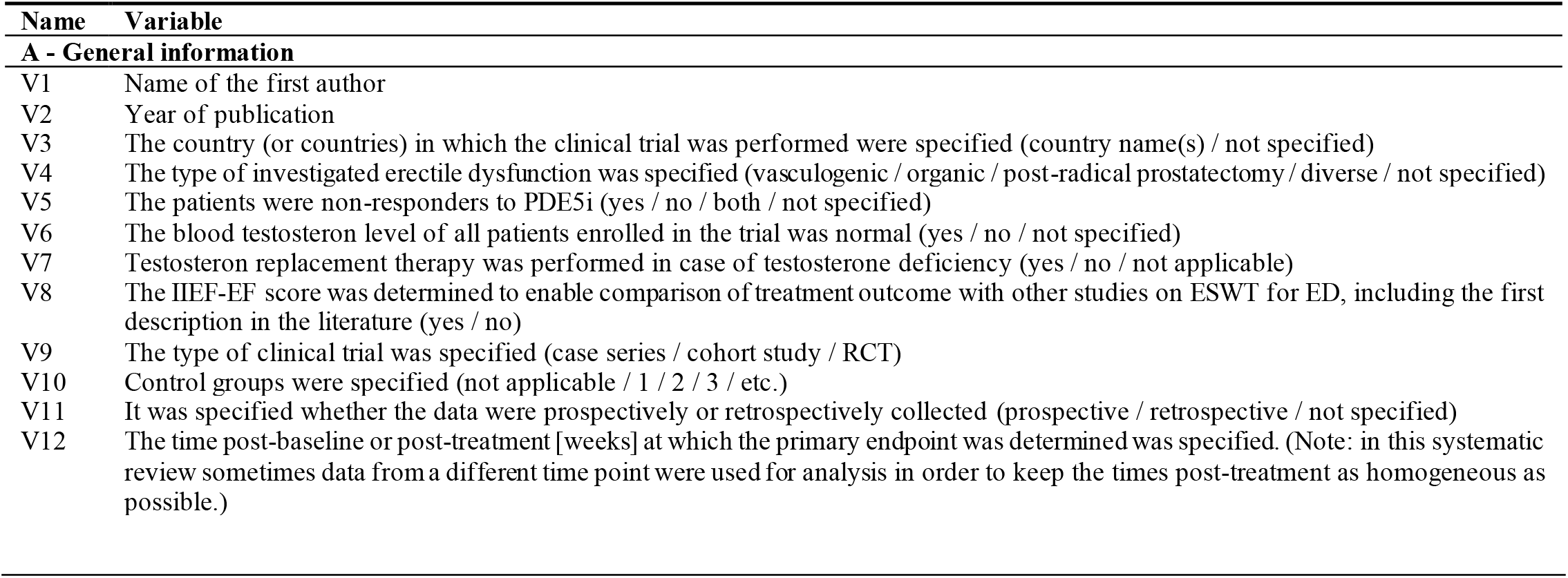
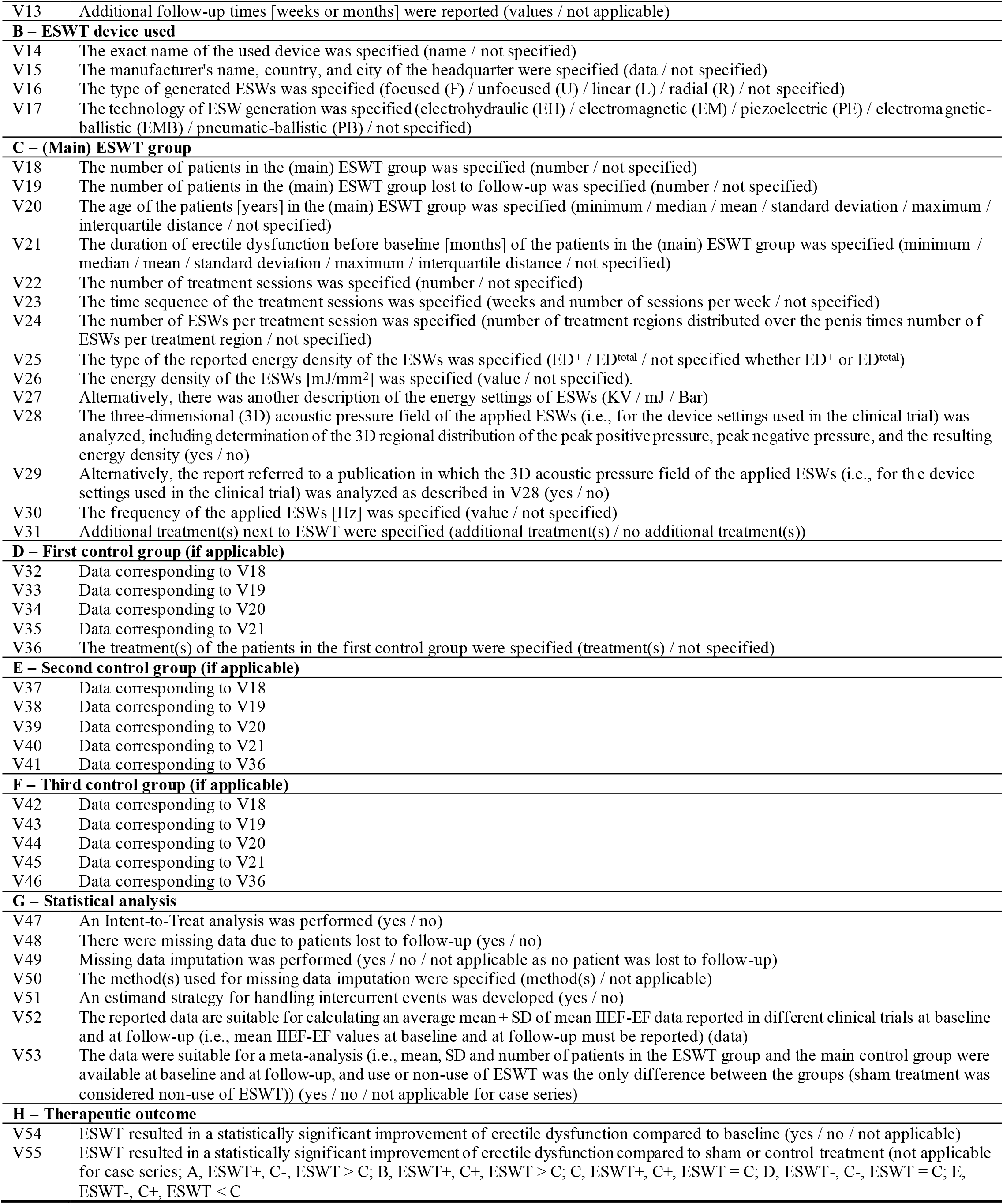
Variables extracted from the 87 reports on extracorporeal shock wave therapy (ESWT) for erectile dysfunction (ED) identified in the systematic literature search outlined in Figure 1.

Statistical analysis (calculation of mean, standard deviation, median and range of the investigated variables as well as Wilcoxon matched-pairs signed rank test for effect estimation) were calculated using GraphPad Prism (Version 10.3.1 for Windows; GraphPad Software, Boston, MA USA). Furthermore, a meta-analysis was performed using the software Comprehensive Meta-Analysis (Version 4; Biostat, Inc., Englewood, NJ, USA). P values smaller than 0.05 were considered statistically significant.

## RESULTS

The 87 eligible studies exhibited considerable heterogeneity in the investigated variables (all extracted values are provided in Supplementary Data).

### Type of ED

In 34 of 87 (39%) studies, vasculogenic ED was investigated, in 18 (21%) studies organic ED, in 12 (14%) studies diverse kinds of ED, in 10 (11%) studies ED post-radical prostatectomy, and in one (1%) study each ED in the presence of Peyronie’s disease, priapism-induced ED, ED on the basis of chronic pelvic pain syndrome, veno-occlusive ED based on hypogonadotropic hypogonadism, post pelvic fractures associated with urethral injury, and multifactorial ED in kidney transplant recipients. In 7 (8%) studies the type of investigated ED was not specified.

### Collection of data

In 62 of 87 (71%) studies, the data were prospectively collected, and in 15 (17%) studies retrospectively. In 10 (11%) studies it was not specified whether the data were prospectively or retrospectively collected.

### Response to PDE5i

In 20 of 87 (23%) studies, the patients were non-responders to PDE5i, whereas in 64 (74%) studies this was not the case. In 3 (3%) studies both responders and non-responders to PDE5i were included.

### Blood testosterone levels

In 34 of 87 (39%) studies, the blood testosterone level of all enrolled patients was normal. In 3 (3%) studies this was not the case and, thus, testosterone replacement therapy was performed in case of testosterone deficiency. However, in 50 (57%) studies the blood testosterone level of the enrolled patients was not specified.

### IIEF-EF score

In 37 of 87 (43%) studies, the International Index of Erectile Function - Erectile Domain (IIEF-EF) score^41^ was determined to enable comparison of treatment outcome with other studies on ESWT for ED, including the first description in the literature.^42^ In 50 (57%) studies this was not the case.

### Number of patients

The number of patients in the (main) ESWT group varied between 5 and 710 (mean, 55; standard deviation (SD), 87; median, 35), and the number of patients in the first (main) control group varied between 10 and 484 (mean, 45; SD, 68, median, 34). In six studies a second control group was investigated in which the number of patients varied between 24 and 178 (mean, 72; SD, 68, median, 34.5). Furthermore, in one study a third control group with 25 patients was investigated.

### Age of the patients

The average age of the patients could not be calculated, as several studies reported only median data. The average of the 64 of 87 (74%) reported mean ages of patients in the (main) ESWT group was 53.3 years ± 8.6 years (mean ± SD), and the average of the 22 (25%) reported median ages of patients in the (main) ESWT group was 58.8 years ± 3.8 years. Furthermore, in 37 (43%) studies the range of the age of the patients in the (main) ESWT group was specified; the lower limit varied between 19 years and 55 years (mean, 36 years; SD, 11 years; median, 33 years) and the upper limit varied between 36 years and 84 years (mean, 71 years; SD, 9 years; median, 72 years). In one study the age of the patients was not specified.

### Duration of ED before baseline

The average duration of ED before baseline of the patients in the (main) ESWT group could neither be calculated, as several studies reported only median data. However, the following information could be extracted from the provided data: (i) in 28 of 77 (36%) studies addressing ED other than post-prostatectomy the mean duration of ED before baseline was specified, with values varying between 5.5 months and 118 months (mean, 43.8 months; SD, 27.7 months; median, 34.6 months); (ii) in 16 of 77 (21%) studies the median duration of ED before baseline was specified; these values varied between 12 months and 68 months (mean, 47.1 months; SD, 15.8 months; median, 46 months); (iii) in 19 of 77 (25%) studies a lower limit of the duration of ED before baseline was reported (3× >3 months, 14× > 6 months and 2× >12 months), and in 1 of 77 (1%) studies an upper limit (<6 months); (iv) in 23 of 77 (30%) studies the range of the duration of ED before baseline was specified; the lower limit varied between 3 months and 36 months (mean, 10.8 months; SD, 7.2 months; median, 9 months) and the upper limit varied between 12 months and 240 months (mean, 116 months; SD, 82 months; median, 84 months); and (v) in 13 of 77 (17%) studies the duration of ED before baseline was not specified.

### ESWT devices used

The electrohydraulic (EH), electromagnetic (EM), piezoelectric (PE) and ballistic principles for generating ESWs are schematically shown in Figure 2 (taken from^11^ which was published under the Creative Commons CC-BY-NC license).

**Figure 2.**
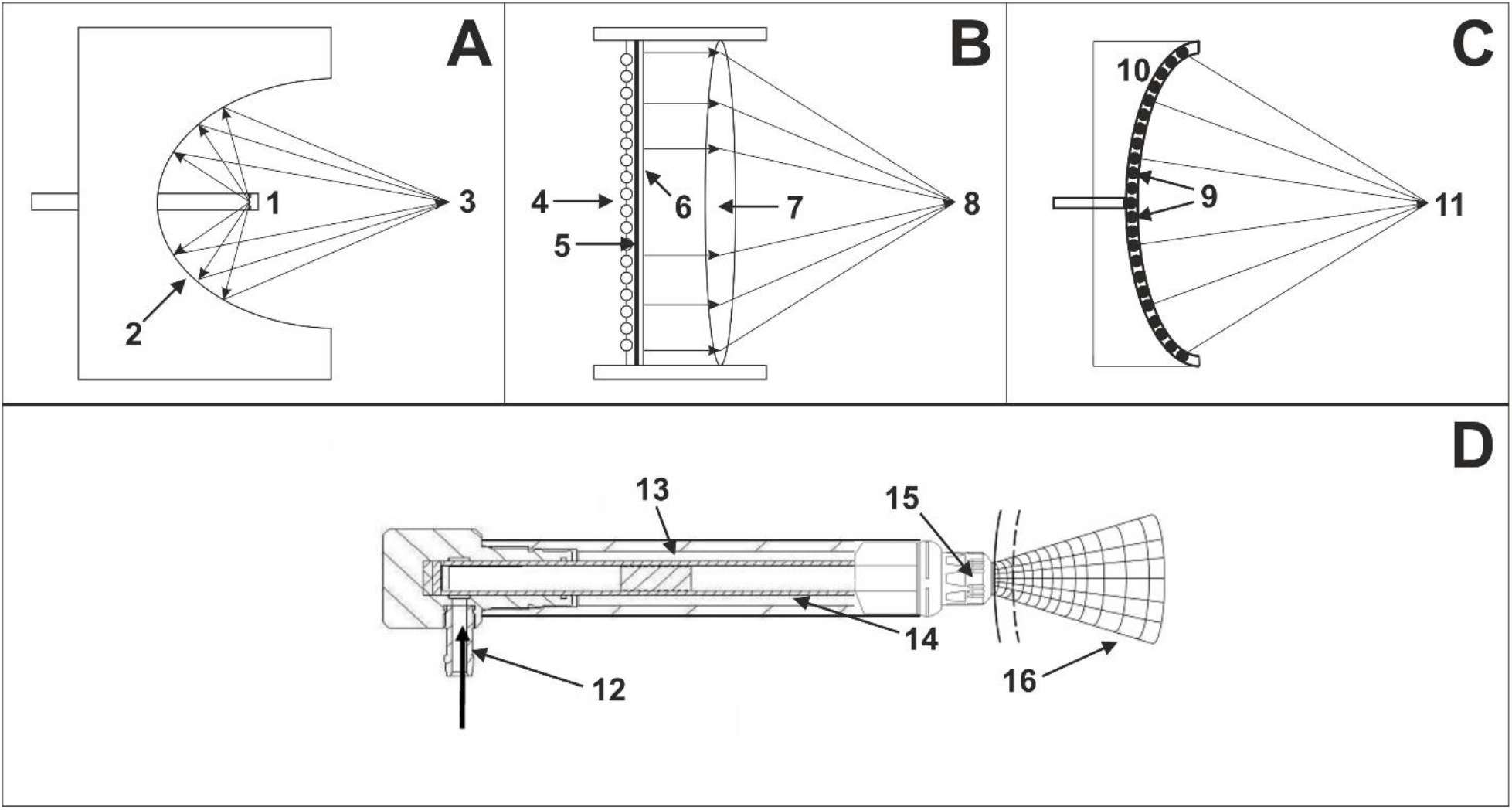
Schematic representation of the mode of operation of focused (A-C) and radial (D) extracorporeal shock wave generators (taken from^11^ which was published under the Creative Commons CC-BY-NC license). (**A**) Electrohydraulic principle(focused extracorporeal shock waves (fESWs)): a high voltage discharges rapidly across two electrode tips (spark-gap) (1) that are positioned in water. The spark-gap serves as the first focal point (1). The heat generated by this process vaporizes the surrounding water. This generates a gas bubble centered on the first focal point, with the gas bubble being filled with water vapor and plasma. The result of the very rapid expansion of this bubble is a sonic pulse, and the subsequent implosion of this bubble causes a reverse pulse, manifesting a shock wave. By means of reflectors of certain shape (2), this shock wave can be converted into a convergent/focused acoustic pressure wave/shock wave with a point of highest pressure at the second focal point (3). ( **B**) Electromagnetic principle (fESWs): a strong, variable magnetic field is generated by passing a high electric current through a coil (4). This causes a high current in an opposed metal membrane (5), which causes an adjacent membrane (6) with surrounding liquid to be forced rapidly away. Because the adja cent membrane is highly conductive, it is forced away so rapidly that the compression of the surrounding liquid generates a shock wave within the liquid. By means of an acoustic lens (7) of certain shape, this shock wave can be converted into a convergent/focused acoustic pressure wave/shoc k wave with a point of highest pressure at a focal point (8). (**C**) Piezoelectric principle (fESWs): a large number of piezocrystals (9) are mounted in a bowl-shaped device (10); the number of piezocrystals can vary from a few to several thousands (typically between 1,000 and 2,000). When applying a rapid electrical discharge, the piezocrystals react with a deformation (contraction and expansion), which is known as the piezoelectric effect. This induces a n acoustic pressure pulse in the surrounding water that can steep into a shock wave. Because of the design of the bowl-shaped device an acoustic pressure wave/shock wave can emerge with a point of highest pressure at a focal point (11). ( **D**) Ballistic principle (rESWT): compressed air (pneumatic principle; 12) or a magnetic field (not shown) is used to fire a projectile (13) within a guiding tube (14) that strikes a me tal applicator (15) placed on the patient’s skin. The projectile generates stress waves in the applicator that transmit pressure waves into tissue (16). Linearly formed extracorporeal shock waves can be generated by using different designs of the components of extracorporeal shock wavegenerators shown in th is figure. For example, in linear piezoelectric devices many subassemblies as the one shown in (C) can be arranged serially.

In 22 of 87 (25%) studies the focused part of the EM device, Duolith SD1 (fESWs; Storz Medical, Tägerwillen, Switzerland) was used, in 18 (21%) studies the EH device, Omnispec ED1000 (fESWs; Medispec, Yehud, Israel), in 10 (12%) studies the EM device, Renova (linear ESWs (lESWs); Direx, Petah Tikva, Israel), in 9 (10%) studies the PE device, Piezowave 2 with FBL10x5G2 handpiece (lESWs; Richard Wolf, Knittlingen, Germany); in 7 (8%) studies the EM device, Aries 2 (fESWs; Dornier MedTech, Weßling, Germany) and in 2 (2%) studies the EH device, UroGold 100 (fESWs; MTS, Konstanz, Germany) Furthermore, in one (1%) study each the following devices were used: BTL-6000 SWT (pneumatic-ballistic (P-B); rESWs; BTL); enPulse Pro (electromagnetic-ballistic (EM-B); rESWs, Zimmer, Neu-Ulm, Germany); ESWO-I 80 mm (EM; fESWs; Shenzhen Hyde Medical Equipment Co, Shenzhen, China); GentlePro (EM-B; rESWs; Zimmer); HB-ESWT-01 (not specified; fESWs; Zhanjiang Haibin Medical Equipment Co, Guangdong, China); Intellect Focus Shockwave Therapy SKU (EM; fESWs; Chattanooga, Chattanooga, TN, USA); LGT-2510B (P-B; rESWs; Guangzhou Longest Medical Technology Co, Guangzhou, China); Masterpuls MP50 (P-B; rESWs, Storz Medical); MoreNova (EM; lESWs; Direx); MT 2000H (EM; fESWs; Urontech, Hwaseong, Korea); OrthoGold 100 (EH; fESWs; MTS, Konstanz, Germany); Piezowave 2 with FB10G6 handpiece (PE; fESWs; Richard Wolf); Swiss DolorClast with EvoBlue Handpiece (P-B; rESWs; Electro Medical Systems) and WellWave (PE; fESWs; Richard Wolf). In 5 (6%) studies the used device was not specified.

It should be mentioned that in one study^43^ in which the Duolith SD1 (Storz Medical) was used a linearly formed transducer head was described, without providing further details. It is unknown whether this linearly formed transducer head generated lESWs similar to the Renova (Direx), MoreNova (Direx) and the Piezowave 2 with FBL10x5G2 handpiece (Richard Wolf), or whether this was a regular fESW transducer head with a standoff with an elongated recess/trough. In the other 21 studies in which the Duolith SD1 (Storz Medical) was used, a linearly formed transducer was not mentioned. As in the same study^43^ the patients were also treated with a curved transducer head (which was probably the regular handpiece of the Duolith SD1 (Storz Medical) that generates fESWs), this study was considered focused ESWT in this systematic review.

### Types of ESWs applied

In 56 of 87 (65%) studies focused ESWs were applied, in 20 (23%) studies linear ESWs, in 6 (7%) studies radial ESWs and in 1 (1%) study unfocused ESWs. In 4 (5%) studies the type of ESWs was not specified or could not be determined by the description of the used device.

### Treatment protocols

The treatment protocols were analyzed both cumulatively and separately for studies in which fESWs, lESWs and rESWs were applied (details of the separate analysis are summarized in Table 3). The most commonly used treatment protocols were Week 1 (W1) to W3 and W7-W9 with one treatment session per week (Protocol A; 16 of 87 (18%) studies), W1-W4 with one treatment session per week (Protocol B; 12 (14%) studies), and W1-W6 with one treatment session per week (Protocol C; also 12 (14%) studies). Various other treatment protocols were used. Of note, Protocols A / B / C were the most commonly used treatment protocol among those studies in which fESWs / lESWs / rESWs were applied.

**Table 3.** Details of the treatment protocols of extracorporeal shock wave therapy for erectile dysfunction used in the 87 clinical trials identified in this systematic review, stratified by the type of the applied extracorporeal shock waves. Data are provided as mean ± standard deviation (median; range). The numbers in brackets indicate the numbers of reports that provided the corresponding raw data.

|  | fESWs | lESWs | rESWs |
| --- | --- | --- | --- |
| TS | 9.0 ± 3.3<br>(9; 4 – 18) [56] | 5.6 ± 2.2<br>(4.5; 4 – 10) [20] | 6.7 ± 1.0<br>(6; 6 – 8) [6] |
| ESWs/TS | 2773 ± 1556<br>(3000; 1500 – 10,000) [55] | 3746 ± 1758<br>(3800; 600 – 6600) [20] | 5000 ± 2828<br>(4500; 2000 – 10,000) [6] |
| ΣESWs | 24,558 ± 16,690<br>(18,000; 8000 – 90,000) [55] | 18,980 ± 9633<br>(18,600; 3600 – 39,600) [20] | 33,000 ± 17,697<br>(27,000; 12,000 – 60,000) [6] |
| EnD [mJ/mm<sup>2</sup>] | 0.14 ± 0.07<br>(0.09; 0.05 – 0.25) [47] | 0.11 ± 0.03<br>(0.09; 0.09 – 0.16) [17] | 0.09 ± 0.0<br>(0.09; 0.09 – 0.09) [2] |
| ΣEnD [J/mm<sup>2</sup>] | 3.25 ± 2.46<br>(1.8; 0.81 – 9) [47] | 2.19 ± 1.44<br>(1.8; 0.54 – 6.34) [17] | 1.89 ± 1.15<br>(1.9; 1.1 – 2.7) [2] |
| F [Hz] | 3.8 ± 1.8<br>(3.7; 1.7 – 8) [44] | 5.3 ± 1.8<br>(5; 2 – 8) [9] | 13.7 ± 2.9<br>(12; 12 – 17) [3] |
| ΣTT [min] | 119 ± 66<br>(116; 33 – 400) [44] | 58 ± 45<br>(48; 20 – 167) [9] | 41 ± 23<br>(33; 24 – 67) [3] |
| P | W1-W3 and W7-W9 (1x per week) [13] | W1-W4 (1x per week) [9] | W1-W6 (1x per week) [3] |

Abbreviations: fESWs, focused extracorporeal shock waves; lESWs, linear extracorporeal shock waves; rESWs, radial extracorporeal shock waves; TS, number of therapy sessions; ESWs/TS, number of extracorporeal shock waves per therapy session; $\Sigma$ ESWs, total number of applied extracorporeal shock waves (TS x ESWs/TS); EnD, energy density; $\Sigma$ EnD, cumulated energy density applied ( $\Sigma$ ESWs x EnD); F, frequency at which the ESWs were applied; $\Sigma$ TT, total treatment time ( $\Sigma$ ESWs / F); P, the most commonly used treatment protocol; W, week.

### Details of the treatment protocols

The number of treatment sessions varied between 4 and 14 (mean, 8.3; SD, 3.3; median, 6; data from 87 studies).

The number of ESWs applied per treatment session varied between 600 and 10,000 (mean, 3022; SD, 1743; median, 3000; data from 86 studies as not in all studies all relevant information was provided).

The total number of applied ESWs varied between 3600 and 90.000 (mean, 23,120; SD, 14,684; median; 18,000; data from 86 studies).

The energy density of the applied ESWs varied between 0.05 Millijoule (mJ)/mm^2^ and 0.25 mJ/mm^2^ (mean, 0.13 mJ/mm^2^; SD, 0.06 mJ/mm^2^; median, 0.09 mJ/mm^2^; data from 71 studies). Obviously incorrect information about the energy density of the applied ESWs (0.009 mJ/mm^2^ as well as 20 mJ/mm^2^, 15 mJ/mm^2^ and 12 mJ/mm^2^)^44-46^ were not considered in these calculations. The same applied to other descriptions of the energy settings of ESWs (including KiloVolt (KV), Millijoule (mJ) and Bar)^47-51^ from which without further information no direct conclusions can be drawn about the energy density of the applied ESWs.

The cumulated energy density applied over all treatment sessions varied between 0.54 J/mm^2^ and 9 J/mm^2^ (mean, 2.83 J/mm^2^; SD, 2.15 J/mm^2^; median, 1.8 J/mm^2^; data from 71 studies).

The frequency of the applied ESWs varied between 1.66 Hz and 17 Hz (mean, 4.3 Hz; SD, 2.7 Hz; median, 4 Hz; data from 58 studies).

From these values the total treatment time was calculated (total number of applied ESWs divided by the frequency at which the ESWs were applied), that varied between 20 min and 500 min (mean, 111 min; SD, 84 min; median, 100 min; data from 60 studies).

Of note, none of the 87 studies specified whether the reported energy density of the ESWs represented the positive or total energy density. Furthermore, no study analyzed the 3D acoustic pressure field of the applied ESWs (i.e., for the device settings used in the study), including determination of the 3D regional distribution of the peak positive pressure, peak negative pressure, and the resulting energy density. Furthermore, none of the studies referenced any publication that provided an analysis of the 3D acoustic pressure field for the ESW device settings used.

### Combination treatments

In 27 of 87 (31%) studies, ESWT was combined with other treatments. These other treatments were PDE5i oral (daily or on demand; 19 studies) and PDE5i oral + L arginine supplement (2 studies), as well as application of ESWs to the pelvic floor, pelvic floor training, transcranial magnetic stimulation, use of a vacuum erectile device, injection of platelet-rich-plasma and subcutaneous injection of recombinant chorionic gonadotropin plus oral intake of Epimedium Breviconum (1 study each). In 60 (69%) studies ESWT was not combined with other treatments.

### Intent-to-Treat analysis

In 51 of 87 (59%) studies, an Intent-to-Treat (ITT) analysis ^52-54^ was performed, whereas in 36 (41%) studies this was not the case. Furthermore, in 38 (44%) studies at least one patient was lost to follow-up in any of the investigated groups, with group-specific lost to follow-up rates varying between 2.3% and 100% (mean, 19.7%; SD, 16.8%; median, 16.7%). However, in only 2 of the 38 (5%) studies with missing data, missing data imputation ^55,56^ was performed (using the Baseline Observation Carried Forward Method).

### Estimand strategy for handling intercurrent events

In only 1 of 87 (1%) studies, an estimand strategy for handling intercurrent events^57-59^ was developed.

### Treatment outcome

Among the 77 studies that did not address ED post-prostatectomy, 71 (92%) studies reported a statistically significant improvement in ED from baseline to follow-up. Furthermore, 33 of these 77 (43%) studies reported mean IIEF-EF values at baseline and at follow-up. The mean IIEF-EF values at baseline varied between 7 and 21.2 (mean, 13.7; SD, 3.4; median, 14) and the mean IIEF-EF values at follow-up varied between 12.3 and 25.8 (mean, 19.5; SD, 3.6; median, 20). Statistical analysis using the Wilcoxon matched-pairs signed rank test demonstrated a significant (p < 0.001) difference in the mean IIEF-EF values at baseline and at follow-up. The mean follow-up time in these 33 studies varied between 7 and 30 weeks (mean, 18.1 weeks; SD, 7.1 weeks; median, 24 weeks).

In 20 of the 47 (43%) cohort studies and RCTs, ESWT was statistically significantly superior to sham/control treatment. Furthermore, in 5 (11%) of these studies ESWT and sham/control treatment significantly improved ED, but ESWT was not significantly superior to sham/control treatment. In only 2 (4%) of these studies neither ESWT nor sham/control treatment were effective, and in none of these studies sham/control treatment was significantly superior to ESWT. For the remaining 20 cohort studies and RCTs the corresponding analysis could not be performed based on the published data.

### Meta-analysis

19 of the 47 (40%) cohort studies and RCTs were suitable for a meta-analysis (i.e., mean and SD of the primary endpoint (e.g., the IIEF-EF score) as well as the number of patients in the (main) ESWT group and the main control group were reported at baseline and at follow-up, and use or non-use of ESWT was the only difference between the groups (sham treatment was considered non-use of ESWT)). The details of the meta-analysis are summarized in Figure 3 and Table 4. For the pooled analysis the standard difference in means was 1.53 (variance; 0.06; lower limit, 1.05; upper limit, 2.00; Z-value, 6.26; p < 0.001).

**Table 4.**
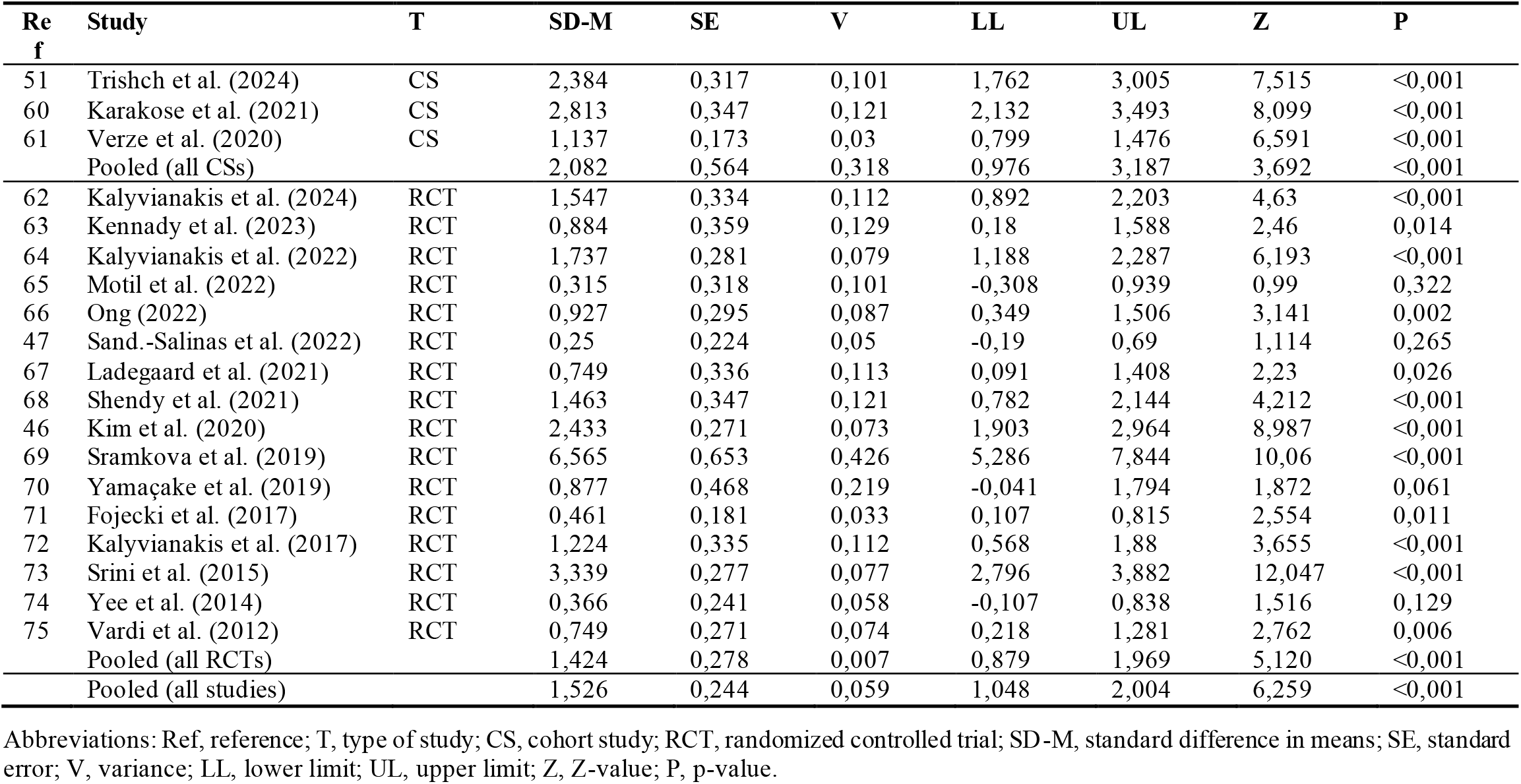
Details of the meta-analysis of 19 cohort studies and RCTs on extracorporeal shock wave therapy for erectile dysfunction performed in this systematic review.

| Ref | Study | T | SD-M | SE | V | LL | UL | Z | P |
| --- | --- | --- | --- | --- | --- | --- | --- | --- | --- |
| 51 | Trishch et al. (2024) | CS | 2,384 | 0,317 | 0,101 | 1,762 | 3,005 | 7,515 | <0,001 |
| 60 | Karakose et al. (2021) | CS | 2,813 | 0,347 | 0,121 | 2,132 | 3,493 | 8,099 | <0,001 |
| 61 | Verze et al. (2020) | CS | 1,137 | 0,173 | 0,03 | 0,799 | 1,476 | 6,591 | <0,001 |
|  | Pooled (all CSs) |  | 2,082 | 0,564 | 0,318 | 0,976 | 3,187 | 3,692 | <0,001 |
| 62 | Kalyvianakis et al. (2024) | RCT | 1,547 | 0,334 | 0,112 | 0,892 | 2,203 | 4,63 | <0,001 |
| 63 | Kennady et al. (2023) | RCT | 0,884 | 0,359 | 0,129 | 0,18 | 1,588 | 2,46 | 0,014 |
| 64 | Kalyvianakis et al. (2022) | RCT | 1,737 | 0,281 | 0,079 | 1,188 | 2,287 | 6,193 | <0,001 |
| 65 | Motil et al. (2022) | RCT | 0,315 | 0,318 | 0,101 | -0,308 | 0,939 | 0,99 | 0,322 |
| 66 | Ong (2022) | RCT | 0,927 | 0,295 | 0,087 | 0,349 | 1,506 | 3,141 | 0,002 |
| 47 | Sand.-Salinas et al. (2022) | RCT | 0,25 | 0,224 | 0,05 | -0,19 | 0,69 | 1,114 | 0,265 |
| 67 | Ladegaard et al. (2021) | RCT | 0,749 | 0,336 | 0,113 | 0,091 | 1,408 | 2,23 | 0,026 |
| 68 | Shendy et al. (2021) | RCT | 1,463 | 0,347 | 0,121 | 0,782 | 2,144 | 4,212 | <0,001 |
| 46 | Kim et al. (2020) | RCT | 2,433 | 0,271 | 0,073 | 1,903 | 2,964 | 8,987 | <0,001 |
| 69 | Sramkova et al. (2019) | RCT | 6,565 | 0,653 | 0,426 | 5,286 | 7,844 | 10,06 | <0,001 |
| 70 | Yamaçake et al. (2019) | RCT | 0,877 | 0,468 | 0,219 | -0,041 | 1,794 | 1,872 | 0,061 |
| 71 | Fojecki et al. (2017) | RCT | 0,461 | 0,181 | 0,033 | 0,107 | 0,815 | 2,554 | 0,011 |
| 72 | Kalyvianakis et al. (2017) | RCT | 1,224 | 0,335 | 0,112 | 0,568 | 1,88 | 3,655 | <0,001 |
| 73 | Srini et al. (2015) | RCT | 3,339 | 0,277 | 0,077 | 2,796 | 3,882 | 12,047 | <0,001 |
| 74 | Yee et al. (2014) | RCT | 0,366 | 0,241 | 0,058 | -0,107 | 0,838 | 1,516 | 0,129 |
| 75 | Vardi et al. (2012) | RCT | 0,749 | 0,271 | 0,074 | 0,218 | 1,281 | 2,762 | 0,006 |
|  | Pooled (all RCTs) |  | 1,424 | 0,278 | 0,007 | 0,879 | 1,969 | 5,120 | <0,001 |
|  | Pooled (all studies) |  | 1,526 | 0,244 | 0,059 | 1,048 | 2,004 | 6,259 | <0,001 |

Abbreviations: Ref, reference; T, type of study; CS, cohort study; RCT, randomized controlled trial; SD-M, standard difference in means; SE, standard error; V, variance; LL, lower limit; UL, upper limit; Z, Z-value; P, p-value.

**Figure 3.**
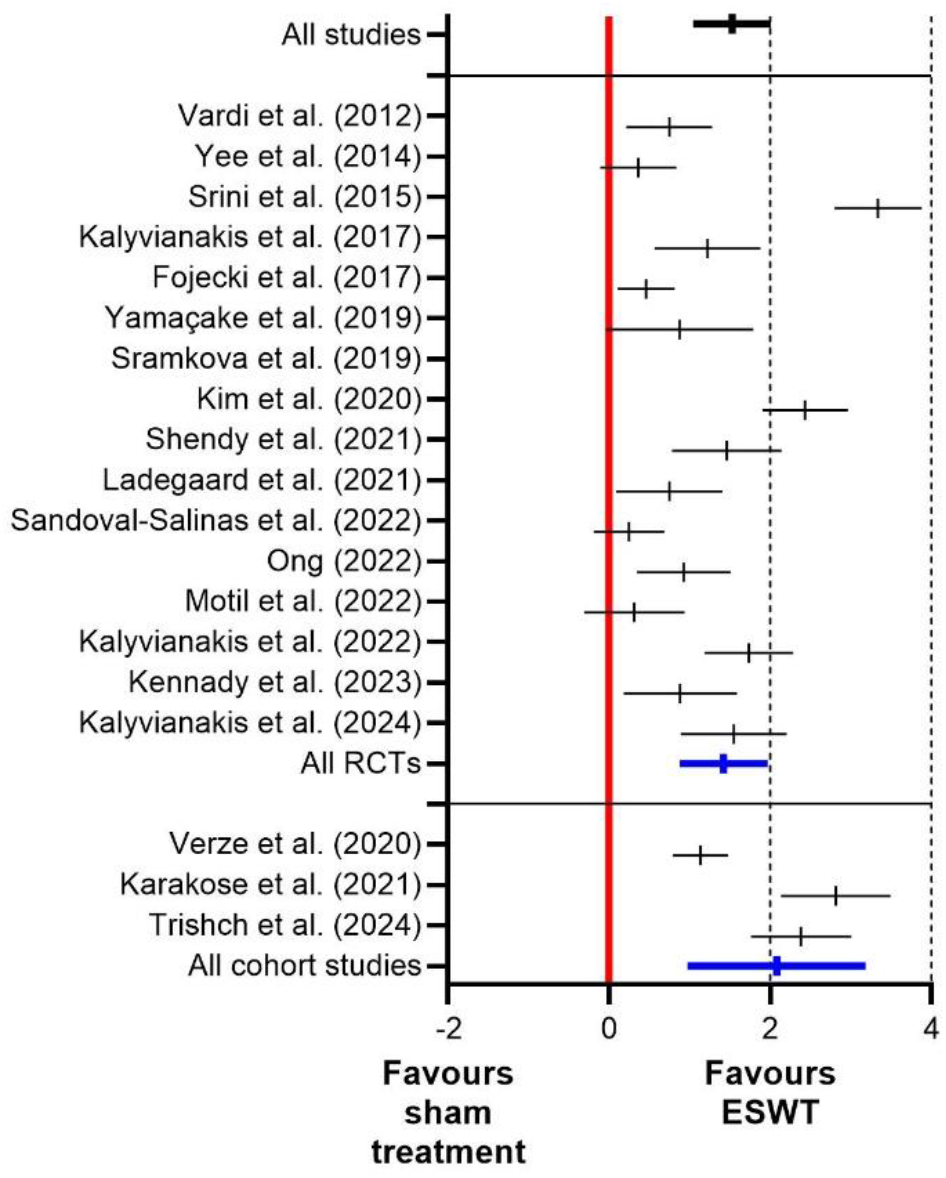
Results of the meta-analysis of 19 cohort studies and randomized controlled trials on extracorporeal shock wave therapy for erectile dysfunction performed in this systematic review.

### Safety

In none of the 87 studies severe adverse events were reported.

## DISCUSSION

The results of this systematic review can be summarized and interpreted as follows: (i) there is clear evidence that ESWT for ED is effective and safe, and can therefore be a valuable treatment modality in the management of ED; (ii) the AUA is correct in its assessment that the true effect of ESWT for ED may be substantially different from the current estimate of the effect; (iii) without a fundamental shift in the planning, implementation and analysis of clinical studies on ESWT in ED, the assessment by the AUA is unlikely to change, and (iv) future studies on ESWT for ED should include adequate characterization and reporting of extracorporeal shock waves, appropriate handling of missing data and intercurrent events, and comprehensive classification of ESWT in the overall context of the available treatment options for ED.

In the following paragraphs each of these aspects is addressed in detail.

### Adequate characterization and reporting of extracorporeal shock waves

Figure 4A was redrawn from an illustration in a recent review on the basic physics of shock waves for ED (^76^; Fig. 2 therein). The curve shown in this figure was described by the authors of^76^ as an idealized pressure vs time plot of a shock wave at one specific point along its trajectory. However, it is important to note that none of the patients suffering from ED in the 87 studies identified in our systematic review were treated with ESWs exhibiting the energy signature shown in Figure 4A. To illustrate this, Figure 4B (taken from^77^ with permission^78^) shows a typical averaged pressure vs time plot (averaged from 25 ESWs) at the focus point (explained below) at the maximum output setting of the Duolith SD1 (Storz Medical) that was used in 22 of the 87 studies identified in our systematic review. Compared to the pressure vs time plot in Figure 4A the corresponding plot in Figure 4B shows (i) a substantially lower peak positive pressure (p^+^) (approximately 43 Megapascal (MPa) vs. 100 MPa), (ii) a steep fall in positive pressure after p^+^ with a steepness similar to the steepness of the rise in positive pressure before reaching p^+^, resulting in a much more symmetric curve around p^+^ than shown in Figure 4A, and (iii) a much longer phase of negative pressure (approximately 2.3 µs vs. approximately 150 ns). The energy density (also outlined below) of the ESWs represented in Figure 4B was determined as 0.23 mJ/mm^2^ by the authors of^77^, whereas the nominal setting of the energy density of the Duolith SD1 (Storz Medical) was 0.55 mJ/mm^2^ during these measurements (c.f. Table 1 in^77^). In line with this finding the authors of^77^ stated that their results compared qualitatively, but not quantitatively with manufacturer specifications.^77^ Moreover, the energy density of the ESWs utilized in ESWT for ED was lower, averaging 0.09 mJ/mm^2^ across the majority of the 87 studies identified in our systematic review. This implies that p^+^ of the ESWs used in ESWT for ED with the Duolith SD1 (Storz Medical) is lower than shown in Figure 4B. According to^77^, ESWs generated by the Duolith SD1 (Storz Medical) with energy density of 0.12 mJ/mm^2^ have a peak positive pressure of 17.5 MPa (Table 1 in^77^), which is much less than the 100 MPa shown in Figure 4A.

**Figure 4.**
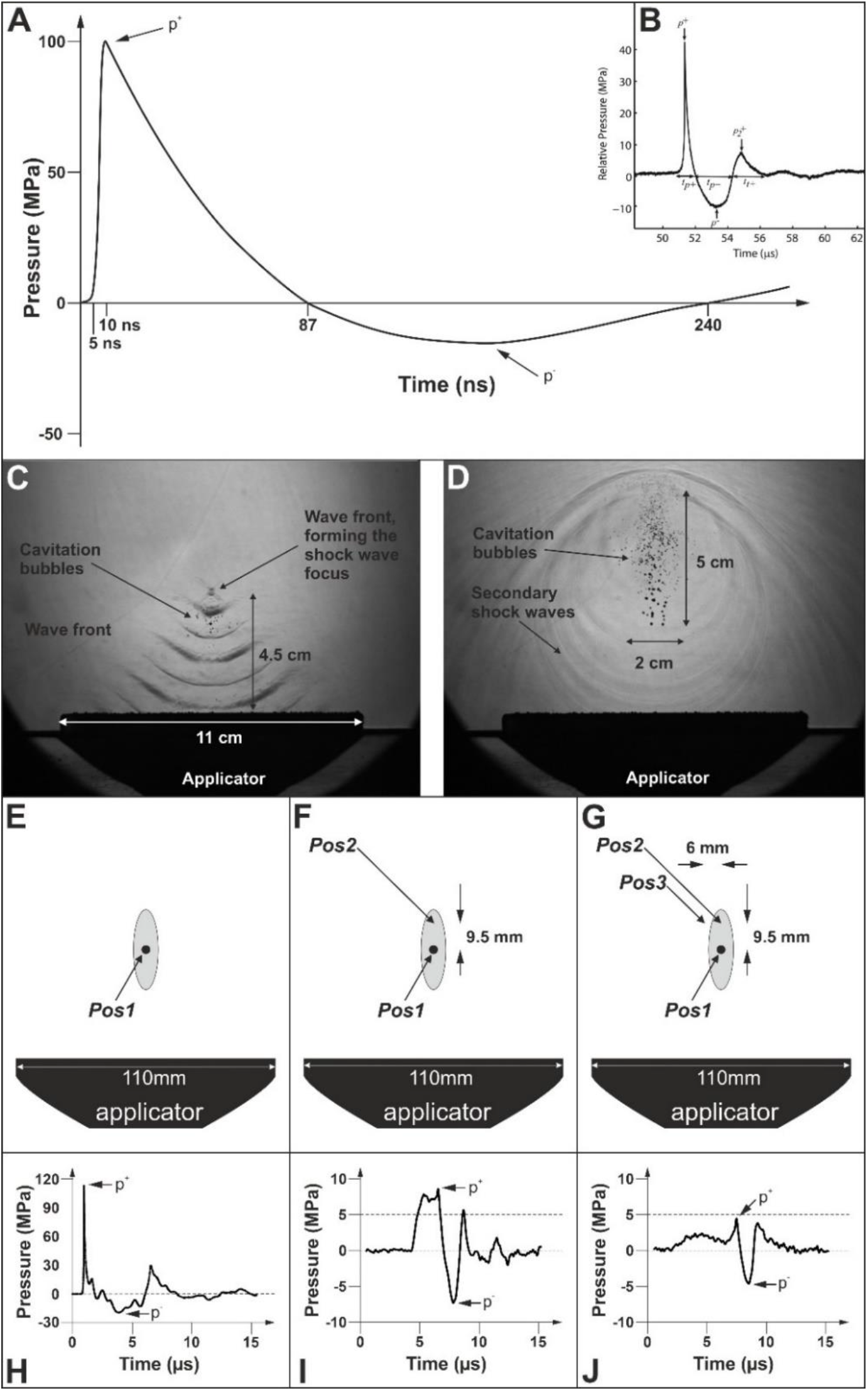
Representative pressure vs time plots and shadow imaging of extracorporeal shock waves for illustration purposes (Part 1). Pa nel B was taken from^77^ with permission^78^; Panels C, D were taken from^80^ which was published under the Creative Commons CC BY 2.0 license; and Panels E-J were taken from^86^ which was published under the Creative Commons CC BY license. Details are in the main text.

Of note, the same study^77^ found that (i) shock formation did not occur for any machine settings of the Duolith SD1 (Storz Medical), and (ii) predictions from simulations suggested that a true shock wave (i.e., a shock wave in a strict physical meaning) could be obtained in water if the initial pressure output of the device were doubled.^77^ Accordingly, the ESWs generated by the Duolith SD1 (Storz Medical) are focused pressure waves rather than focused true shock waves. This is important because (at least in the context of using the Duolith SD1 (Storz Medical)) there is no scientific justification for distinguishing between focused shock waves and radial pressure waves in ESWT for ED (as done in, e.g.,^47,76^), despite the fact that focused ESWs and radial ESWs have different energy signatures.^11,79-81^ Importantly, the question of whether an acoustic wave is a shock wave or a pressure wave is only decided by the part with positive pressure, but not by the part with negative pressure. The findings in^77^ raise the question of whether all ESWT treatments of ED are actually performed with focused pressure waves rather than with focused true shock waves. The information provided in the corresponding review articles^76,82^ is not suitable to answer this question. In reality, the matter is even more complicated: even if an ESWT device would be used in ESWT for ED that could generate focused true shock waves, and this device would be operated such that it would actually generate focused true shock waves, this would not imply that the entire target tissue would be treated with focused true shock waves. This is due to the fact that the 3D acoustic pressure fields generated by ESWT devices are not homogeneous with respect to the maximum pressure, energy density and other characteristics of the ESWs.

To illustrate this, Figure 4C, D shows a two-dimensional (2D) representation of the 3D, rotationally symmetric, acousticpressure field generated by an ESWT device that has not been used in the management of ED (PiezoClast; Electro Medical Systems, Nyon, Switzerland). These pictures (taken from^80^ which was published under the Creative Commons CC BY 2.0 license) were generated using shadow imaging which requires a powerful light source and a high-speed camera (c.f.^83^). Positive pressure in water results in disturbances that can refract light rays, so they can cast shadows (c.f. Figure 4C). The same applies to cavitation bubbles that occur during the phase of negative pressure of ESWs (Figure 4D).^83,84^ In general, cavitation bubbles occur when the pressure in a liquid quickly drops below the vapor pressure.^85^ When these cavitation bubbles collapse at the end of the phase of negative pressure of an ESW, a small, secondary shock wave is generated that is a true shock wave in a strict physical meaning. The merging of many of these secondary shock waves, triggered by the almost simultaneous collapse of many cavitation bubbles, can then lead to larger secondary shock waves that propagate radially (as shown in Figure 4D). The elliptical field of cavitation bubbles in Figure 4D already indicates the absence of a homogeneous acoustic pressure field above the applicator of the ESWT device used.

Figure 4E-G (modified from^86^ which was published under the Creative Commons CC BY license) shows graphical representations of the applicator of the same ESWT device that was used for shadow imaging in Figure 4C, D (PiezoClast; Electro Medical Systems). Position 1 (indicated by a black point) at 4.5 cm above the applicator represents the spot in Figure 4C where the wave front formed the shock wave focus. This point is also named the focus point. Figure 4H shows the pressure vs time plot of the ESWs measured at this focus point (Figure 4H-J was modified from^86^ which was published under the Creative Commons CC BY license). The pressure vs time plot shown in Figure 4H has a peak positive pressure (p^+^) of approximately +110 MPa (exceeding the threshold of +100 MPa indicated in Figure 4A) and a peak negative pressure (p^-^) of approximately -20 MPa; both values exceed the corresponding values in Figure 4B. The energy density of the corresponding ESWs was determined as 0.4 mJ/mm^2^ in^86^. Furthermore, the pressure vs time plot shown in Figure 4H fulfills the requirements set out in^87^ of a focused true shock wave. Figure 4I shows the pressure vs time plot of the same ESWs measured at Position 2 in Figure 4F, which was 9.5 mm above the focus point in the 3D acoustic pressure field. One can see that the absolute values of p^+^ and p^-^ were substantially lower at Position 2 (p^+^ = +8.7 MPa; p^-^ = -7.5 MPa) than at the focus point (p^+^ ∼ +110 MPa; p^-^ ∼ -20 MPa). Most importantly, the pressure vs time plot measured at Position 2 was not simply a scaled-down version of the pressure vs time plot measured at the focus point but showed a different steepness of the rise in positive pressure before reaching p^+^. More generally speaking, unlike the situation at the focus point (c.f. Figure 4H) the pressure vs time plot measured at Position 2 (Figure 4I) did not fulfill the requirements set out in^87^ of a focused true shock wave.

Figure 4J shows the pressure vs time plot of the same ESWs measured at Position 3 in Figure 4G, which was 9.5 mm above and 6 mm left to the focus point in the 3D acoustic pressure field. At this position p^+^ did not even reach +5 MPa, and the pressure vs time plot measured at Position 3 was neither a scaled-down version of the pressure vs time plot measured at the focus point nor measured at Position 2. Of note, the ratio of the absolute values of p^+^ and p^-^ was approximately 5.5 (110 MPa /20 MPa) at the focus point, 1.16 (8.7 MPa / 7.5 MPa) at Position 2 and 0.91 (4.3 MPa / 4.8 MPa) at Position 3.

These measurements demonstrate different acoustic pressure conditions (and, thus, different energy densities) within the 3D acoustic pressure field of an ESW. One characteristic of this 3D acoustic pressure field is the -6 dB (or -3 dB) focus, which is the 3D region around the focus point at which the local p^+^ is at least 50% (or at least 70.8%) of p^+^ at the focus point.^79^ Another characteristic is the 5 MPa focus, which is the 3D region around the focus point at which the local p^+^ is at least 5 MPa, regardless of p^+^ at the focus point.^79^ For the ESWs whose pressure vs time plot at the focus point is shown in Figure 4H, the 5 MPa focus is indicated as gray ellipsoids in Figure 4E-G. Position 2 was inside the 5 MPa focus whereas Position 3 was not.

The energy density of an ESW is defined as

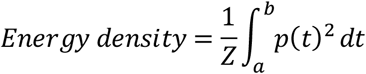

with Z the impedance of sound in water (1.5×10^6^ kg/m^2^s), p(t) the pressure as a function of time and the integration limits a and b. According to IEC-61846^88^ there are two types of integration limits. The positive temporal integration limits are defined as the times between which the positive pressure first exceeds 10% of p^+^ (a^+^ in Figure 5A,B) and the first time it reduces below 10% of p^+^ (b^+^ in Figure 5A,B)^88^ (Figure 5A-D was modified from^86^ which was published under the Creative Commons CC BY license). Furthermore, the total temporal integration limits are defined as the times between which the absolute value of the pressure pulse waveform first exceeds 10% of p^+^ (a^total^ in Figure 5C,D) and the last time it reduces below 10 % of p^+^ (b^total^ in Figure 5C,D).^88^ To simplify matters, the hatched areas under the pressure vs time plots in Figure 5A,B represent the positive energy density, and the hatched areas under the pressure vs time plots in Figure 5C,D the total energy density.

**Figure 5.**
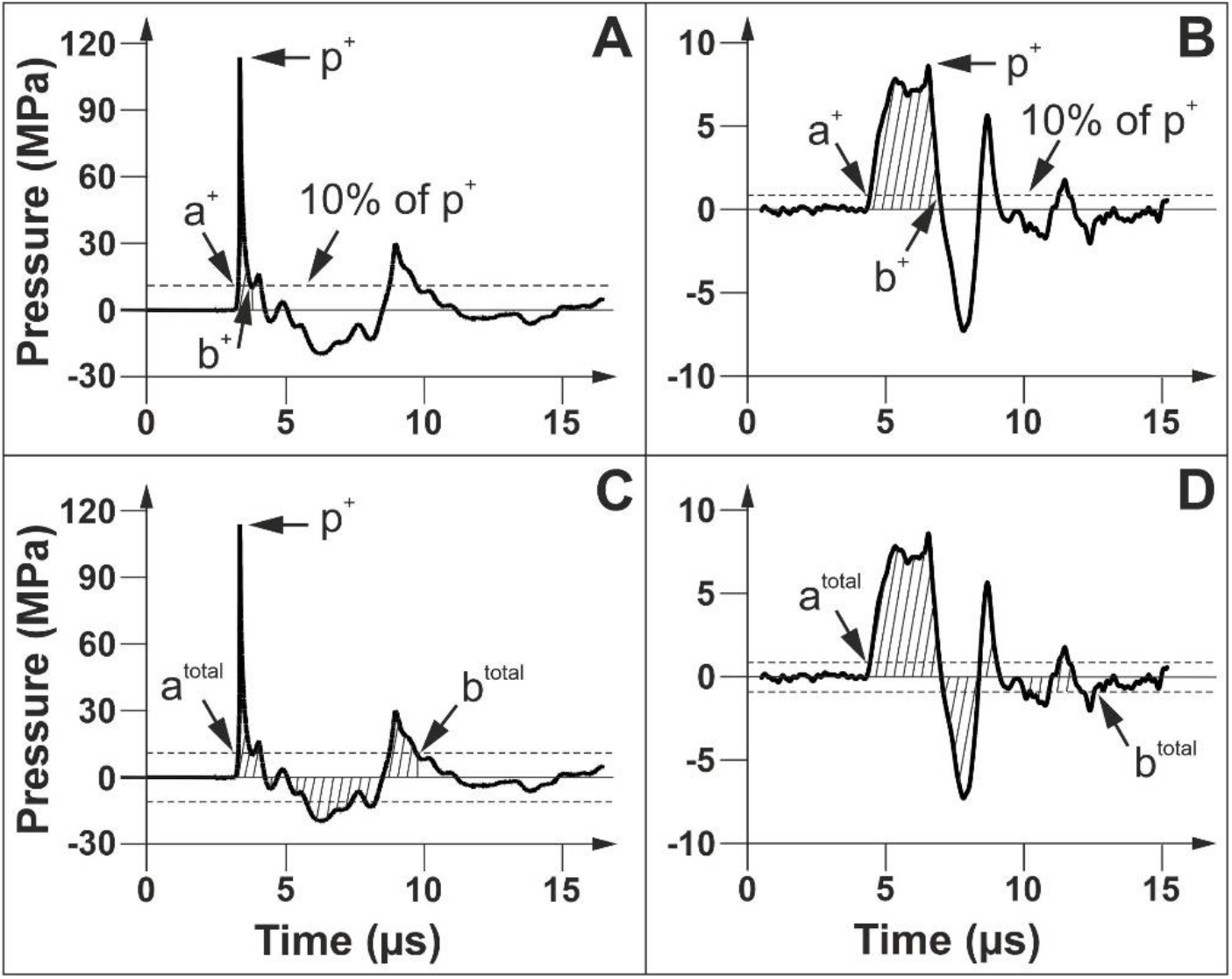
Representative pressure vs time plots of extracorporeal shock waves for illustration purposes (Part 2). The curves in ( **A**,**C**) are the same as in Figure 3H, and the curves in (**B**,**D**) are the same as in Figure 3I. Panels A-D were taken from^86^ which was published under the Creative Commons CC BY license. Details are in the main text.

Accordingly, the energy density within the 3D acoustic pressure field of an ESW exhibits substantial regional variation. Publications on ESWT for ED typically report the energy density at the focus point. However, none of the 87 studies on ESWT for ED identified in our systematic review stated whether the reported energy density values were the positive or the total energy density. It is important to understand that without this information, none of these 87 studies on ESWT for ED can be independently reproduced.

At first glance one might be inclined to simply assume that the total energy density is a certain multiple of the positive energy density. However, due to complex nonlinear wave physics, ESWs generated by certain ESWT devices may exhibit markedly different energy signatures. For example, Figure 6 shows 2D representations of the positive pressure, negative pressure and energy density of fESWs generated by the Duolith SD1 (Storz Medical) operated at highest settings and with its short standoff (15 mm) obtained by modeling (the panels were taken and modified from^77^ with permission^78^, and were squeezed / expanded to scale in X and Y to match the drawing of the -3 dB focus zone of the corresponding ESWs in the same publication^77^, which is shown in Figure 6A). One can see that the spatial distribution of positive pressure (Figure 6B) formed an ellipsoid with highest pressure at the position of the focus point (white arrow in Figure 6B), whereas the spatial distribution of negative pressure (Figure 6C) showed the highest absolute value (gray arrow in Figure 6C) at a position that was approximately 18 mm closer to the applicator than the focus point (white arrow in Figure 6C). As a result, the spatial distribution of the total energy density was also shifted towards the applicator, and the maximum total energy density (gray arrow in Figure 6D) was not found at the position of the focus point (white arrow in Figure 6D). As mentioned above, the spatial distributions of the positive pressure, negative pressure and energy density shown in Figure 6 do not represent the situation in ESWT for ED using the Duolith SD1 (Storz Medical), as this device is not operated at highest settings in ESWT for ED. To our knowledge comprehensive information corresponding to the one provided in Figure 6B-D has not been published for any other ESWT device used in ESWT for ED.

**Figure 6.**
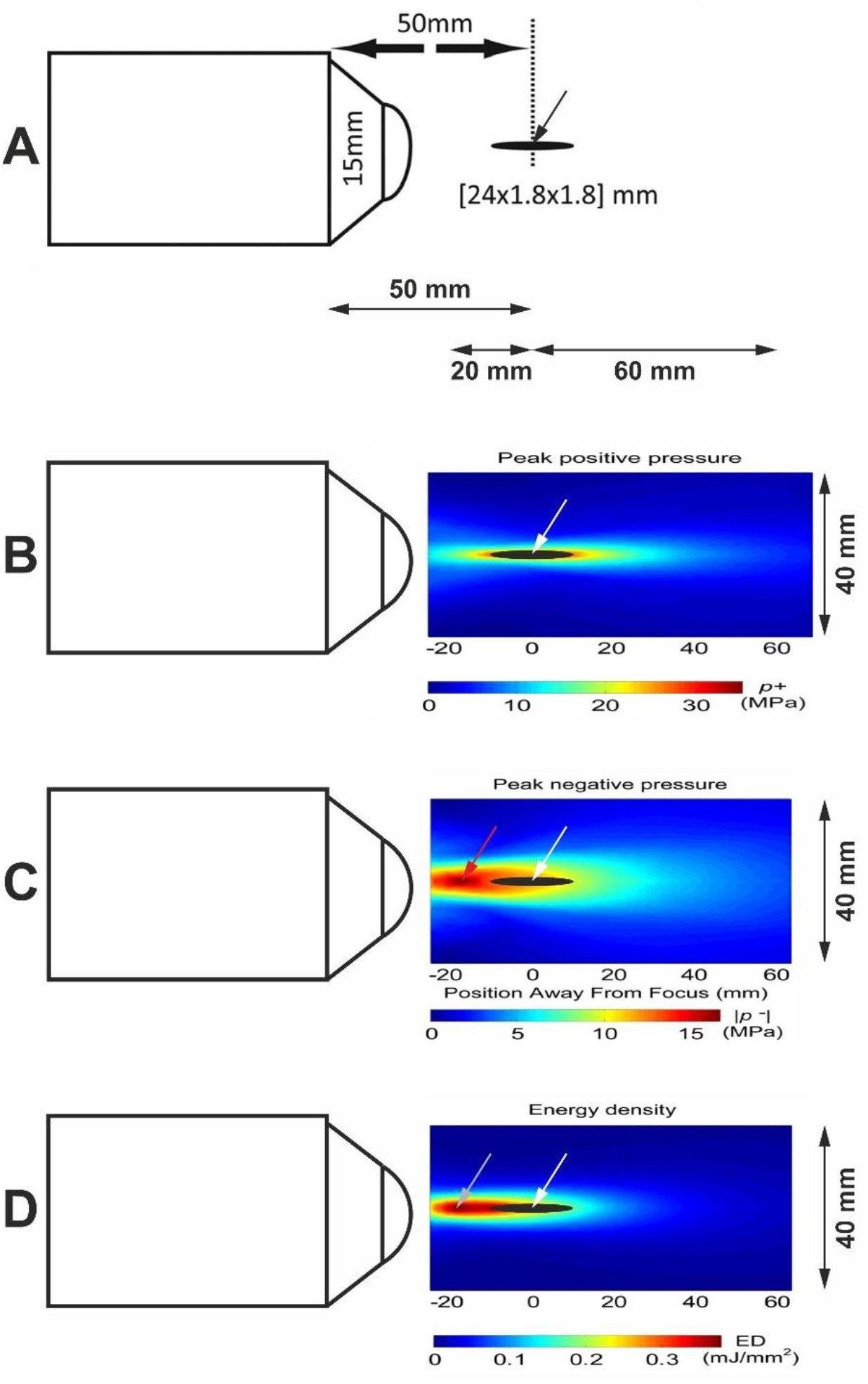
Two-dimensional (2D) representations of the spatial distribution of the positive pressure, negative pressure, and energy density of ESWs generated by the Duolith SD1 (Storz Medical) operated at highest settings and with its short standoff obtained by modeling. Panels A-D were taken and modified from Figures 1 and 11 in^77^ with permission^78^. Details are in the main text.

What implications do these findings have for cells in the target tissue? Cells exposed to ESWs may react differently to the shear stress induced by the positive pressure, as well as to the formation and collapse of cavitation bubbles caused by the negative pressure.^77,83,84^ To our knowledge, only three studies addressed the potential impact of cavitation (and, thus, the negative pressure of ESWs) on cells in vitro and biological tissue under experimental conditions.^89-91^ Specifically, human fetal foreskin fibroblasts in vitro,^89^ *C. elegans* worms^90^ and explanted frog sciatic nerves^91^ were exposed to ESWs either in water / saline or in polyvinyl alcohol (PVA), a high viscosity solution with low cavitation activity during ESW exposure, an acoustic impedance that is nearly identical to that of water, and relatively low ESW attenuation.^92^ In all of these experiments a substantial contribution of cavitation to the effects of ESWs on cells and biological tissue was demonstrated.^89-91^

Considering the small size of cells (with diameters between 10 and 50 µm) relative to the size of ESWs (Figure 4C), it seems implausible that cells can discern whether they are hit by a focused ESW or a radial ESW. However, cells may react differently to various energy signatures of ESWs, including differences in p^+^ and p^-^, duration of exposure to positive and negative pressure, energy density, and the contribution of positive and negative pressure to the total energy density. Accordingly, cells in the target tissue may react very differently to exposure to ESWs, depending on their position within the 3D acoustic pressure field of the applied ESWs.

The specification of respectively KV values (as in^48^), mJ values (as in^49,51^) or bar values (as in^47,50^) does not allow to draw conclusions about the characteristics of ESWs without additional information. This is due to the fact that these values describe the input energy in different types of ESWT devices to generate ESWs rather than the energy density of the resulting ESWs. Specifically, KV (Kilovolt) describes the current that is rapidly discharged across two electrode tips (spark-gap) to vaporize the surrounding water in electrohydraulic ESWT devices (c.f. Figure 2A).^11^ For the EH device, Evotron (HealthTronics, Marietta, GA, USA; not used in any of the 87 studies identified in our systematic review) it was shown that different energy settings (and, thus, different KV values) resulted in ESWs with very similar energy density.^84^ Furthermore, mJ describes the energy that is used to electromagnetically accelerate the bullet in certain radial ESWT devices (c.f. Figure 2D), including the enPulse pro device (Zimmer, Neu-Ulm, Germany; used in^49^ for treating ED). It would be necessary to determine the energy density of the resulting rESWs as done for pneumatically generated rESWs.^83,93^ Moreover, the term Bar describes the air pressure that is used to accelerate the bullet in pneumatic, radial ESWT devices (c.f. Figure 2D). For the Swiss DolorClast with the EvoBlue handpiece and 15 mm applicator (Electro Medical Systems; used in^87^ for treating ED) a good correlation between the air pressure and the energy density of the resulting rESWs was demonstrated, with a loss of approximately 20% in the positive energy density between rESWs generated at 1 Hz and those generated at 25 Hz.^93^ In contrast, the radial ESWs generated by the MasterPuls 200 Ultra with Falcon handpiece and 15 mm applicator (Storz Medical; not used in any of the 87 studies identified in our systematic review) showed a loss of up to 90% in the positive energy density between rESWs generated at 1 Hz and those generated at 21 Hz.^93^ Furthermore, at 1 Hz frequency the rESWs generated by the MasterPuls 200 Ultra / Falcon handpiece / 15 mm applicator (Storz Medical) had consistently lower positive energy densities than the rESWs generated by the Swiss DolorClast / EvoBlue handpiece / 15 mm applicator (Electro Medical Systems).^93^ To our knowledge the corresponding characteristics of the rrESWs generated by the BTL-6000 SWT (BTL; used in^50^ for treating ED) and the MasterPuls 50 (Storz Medical; used in^47^ for treating ED) have not been published.

Finally, it should be noted that severalstudies on ESWT for ED either lacked information on the energy density and other characteristic of the ESWs used,^94-98^, or includedinformation that was evidently inaccurate.^44,46^ Specifically, in^44^ it was stated that patients received 6 treatment sessions, with 3 x 1000 focused ESWs (Duolith SD1; Storz Medical) with energy densities of 12 mJ/mm^2^, 15 mJ/mm^2^ and 20 mJ/mm^2^. Furthermore, in^46^ it was stated that patients received 12 treatment sessions, with 3 x 1000 focused ESWs (MT 2000H device; Urontech Korea, Hwaseong, Korea) with energy densities of 12 mJ/mm^2^, 15 mJ/mm^2^ and 20 mJ/mm^2^. However, these values neither represent the energy density of single fESWs nor the cumulated energy density of all fESWs applied to a certain penile region or during a treatment session. Moreover, in^45^ it was stated that patients received 12 treatment sessions, with 5 x 300 fESWs (Aries 2; Dornier MedTech, Weßling, Germany) with energy density of 0.009 mJ/mm^2^. This is only 10% of the 0.09 mJ/mm^2^ used in the majority of the 87 studies identified in our systematic review and most likely a typographical error. In summary, the characteristics of the 3D acoustic pressure fields to which penile tissue is exposed during ESWT for ED remain largely unknown. In particular, different ESWT devices that are operated at the same energy density at the focus point (e.g., 0.09 mJ/mm^2^) may expose penile tissue to very different 3D acoustic pressure fields. There is no doubt that the precise characterization of these 3D acoustic pressure fields is a mandatory prerequisite for optimizing treatment protocols to improve the effectiveness of ESWT for ED. We hypothesize, over time, this improved assessment of ESWT for ED may contribute to a more favorable assessment by organizations such as the AUA. It is clear that manufacturers of ESWT devices play a critical role in the characterization of the 3D acoustic pressure fields of the ESWs generated by their devices under exactly the conditions used in ESWT for ED.

### Appropriate handling of missing data and intercurrent events

Consider a hypothetical clinical study investigating the efficacy of three novel drugs for treating ED. In this study, researchers randomly assigned n=150 patients suffering from vasculogenic ED to three groups (Groups A, B and C; n=50 patients each). Each group received one of three drugs (Drugs A, B and C). The patients’ IIEF-EF score was determined at baseline and at six months post-treatment initiation (M6). At baseline, all groups had an average IIEF-EF score of 8 ± 2 (mean ± SD), indicating that the patients suffered from severe ED. At M6, n=30 / 45 / 40 patients in Groups A / B / C could be analyzed and had an average IIEF-EF score of 26 ± 3 / 12 ± 4 / 20 ± 4. Statistical analysis of the results with analysis of variance (ANOVA) followed by Bonferroni post tests for pairwise comparison demonstrated statistically significant differences in the mean IIEF-EF score at M6, with Group A > Group C > Group B. Based on these results, the investigators concluded that Drug A is superior to Drugs B and C, while Drug B is superior to Drug C in treating ED. Consequently, they recommended Drug A as preferred option in the management of ED.

Later the following was revealed. In Group A, n=20 patients suddenly and unexpectedly died between baseline and M6, and all deaths were probably or possibly related to study treatment. In Group B, n=5 patients moved to another country shortly after the start of the treatment and were lost to follow-up. In Group C, the patients quickly realized that Drug C was not effective and may be a placebo treatment. As a result, n=10 patients in Group C dropped out of the study because of lack of efficacy, and n=30 patients in Group C used concomitant medication which was not disclosed to the study investigators. Considering this additional information, Drug A must not be used in the management of ED, Drug B was not effective, and the real impact of Drug C on improvement of ED could not be determined.

This admittedly extreme example of a hypothetical clinical study and its results demonstrates the absolute necessity and clinical relevance of critical missing and non-missing data imputation in clinical studies. In this example, missing data imputation would have been necessary in all groups. However, the reasons for missing data were completely different among the groups, which requires different strategies and methods for missing data imputation. Furthermore, in Group C the data obtained at M6 of those patients who used undisclosed concomitant medication were biased and were to be replaced, which requires non-missing data imputation.

More broadly, clinical study results for ED should be subjected to an ITT analysis, with estimand strategies for handling intercurrent events developed and clearly documented in the study protocol. A detailed discussion of these analyzes and strategies lies beyond the scope of this review, as they are thoroughly addressed elsewhere.^52-59^

In brief, ITT analysis includes all patients in a clinical study, regardless of whether the patients completed the study.^52-54^ All leading methods for assessing the methodological quality of RCTs include a critical check of whether an ITT analysis was performed.^99-104^ Missing data can be Missing Completely at Random (which means that there is no relationship between the missingness of the data and any values, observed or missing), Missing at Random (which implies a systematic relationship between the propensity of missing values and the observed data, but not the missing data) or Missing Not at Random (which implies a relationship between the propensity of a value to be missing and its values), respectively.^55^ Various methods are available to handle missing data in results of clinical studies, including Baseline Observation Carried Forward, Last Observation Carried Forward, the Expectation Maximization Algorithm, Treatment based Monte Carlo Markov Chain multiple imputation, among others.^55,56^

The four components of an estimand are Population (i.e., the target population for the research question), Variables (i.e., the endpoints that were obtained from all patients), Intercurrent Events (i.e., all events that occurred after treatment initiation and either precluded the observation of a variable, or affected its interpretation) and Population-Level Summary (i.e., the variables on which the comparison between treatments was based).^57-59^ Furthermore, there are five strategies how to handle intercurrent events: Treatment Policy strategy (in this case it is irrelevant whether an intercurrent event has occurred or not; the data will be collected and analyzed regardless), Composite Strategy (in this case the occurrence of the intercurrent event is incorporated into the endpoint), Hypothetical Strategy (in this case the aim of the clinical study is to estimate the treatment effect in the hypothetical situation where the intercurrent event did not occur), Principal Stratum Strategy (in this case the aim is to estimate the treatment effect within the stratum of patients for whom the intercurrent event did not occur) and Wile on Treatment Strategy (in this case measurements of the endpoint are considered up until the time of the intercurrent event).^57,58,105^

A detailed assessment of the 87 studies on ESWT for ED included in our systematic review, specifically regarding ITT analysis and development of estimand strategies with focus on handling intercurrent events, revealed concerning findings. No study included a detailed strategy for handling intercurrent events as outlined above, and the term Estimand was absent from all of these studies. In one study^44^ rudimentary intercurrent events handling was performed by excluding data of patients from the analysis who reported de novo use of erectogenic aids at the first follow-up. Furthermore, in 38 (44%) studies at least one patient was lost to follow-up in any of the investigated groups, but in only two of these studies^106,107^ missing data imputation was performed (note that the absence of missing data should not be interpreted as a justification for not developing a detailed strategy for handling intercurrent events, as the latter also includes potential non-missing data imputation). Considering that at least 15 studies identified in our systematic review described retrospective analysis of existing data (in 10 studies it was not described whether the data were prospectively or retrospectively collected) the relative number of prospective studies with missing data was even higher. Meta-analyses may be particularly sensitive to lack of missing data imputation. In the case of the meta-analysis performed in this review, in 8 of 19 (42%) studies data were missing that were not imputed.

In the following section, three studies identified in our systematic review are shortly described as examples, in which missing data imputation was either not or incorrectly performed. (i) In^108^ n=710 patients suffering from ED were retrospectively analyzed after five weekly ESWT sessions. The last follow-up examination took place one month after the last treatment (i.e., nine weeks after baseline; W9). At this time 298 of 710 (42%) patients were lost and only 412 patients were examined at W9.^108^ Thus, the results reported at W9 represented the results of only 58% of the patients who were included in the study but were compared with the results obtained from all 710 patients after the last treatment.^108^ No attempt was made to impute the missing data as a prerequisite for performing an ITT analysis. (ii) In^109^ n=115 patients were non-randomly assigned after radical prostatectomy to treatment with PDE5i (n=59) or ESWT + PDE5i (n=56). The last follow-up examination took place six months after surgery (M6). At this time 15 of 56 (27%) patients in the ESWT + PDE5i group and 20 of 59 (34%) patients in the PDE5i group were lost to follow-up. Again, no attempt was made to impute the missing data as a prerequisite for performing an ITT analysis. (iii) In^106^ n=66 patients suffering from ED were randomly assigned to ESWT (n=44) or placebo treatment (PT; n=22). Follow-up examinations were performed at three months after baseline (M3), M6 and M12. However, only patients who were considered treatment responders were examined at M6 and M12. The number of examined patients in the ESWT / PT groups were 42 / 22 at M3, 26 / 4 at M6 and 19 / 0 at M12. Accordingly, only 19 of 44 (43%) of the patients in the ESWT group and 0 of 22 (0%) of the patients in the PT group were considered treatment responders and examined at M12. Missing data (i.e., data of patients who were not considered treatment responders and, thus, not examined at M3, M6 and/or M12) were imputed using the Baseline Observation Carried Forward method. Due to this approach the results of the PT group at M12 were exactly the same as the data at baseline, although no patient in the PT group was examined at M12.

In summary, the almost complete lack of developing strategies for handling intercurrent events as well as missing or incorrect ITT analysis in approximately one third of the 87 studies on ESWT for ED identified in our systematic review should be considered a major obstacle to determining the actual effect of this treatment modality in the management of ED. The current assessment of ESWT for ED by organizations such as the AUA (investigational; Evidence Level Grade C) may be directly related to this issue. While managing missing data and intercurrent events in clinical study analyses can be complex, the literature referenced in this review^52-59^ offers a valuable introduction to the relevant concepts and methods. Furthermore, missing data imputation can be accessed through statistical software such as R^110^ that is also available for free.^111^

It should be mentioned that long-term monitoring of the efficacy of ESWT for ED in comparison with other treatment modalities (including sham/placebo ESWT) could greatly benefit from analysis using the Kaplan-Meier estimator.^112^ Briefly, in oncology research the Kaplan-Meier estimator is widely used to measure the fraction of patients living for a certain amount of time after treatment. This can easily be modified to measure the fraction of patients exceeding, e.g., a certain IIEF-EF score for a certain amount of time after treatment, similar to the use of the Kaplan-Meier estimator in determining long-term outcome after treatment of knee osteoarthitis.^113-115^

### Classification of ESWT in the overall context of the available treatment options for ED

The search for optimal combinations of ESWT with other treatment modalities in the management of ED is currently still in its infancy. In a recent, comprehensive review^3^ the following treatment modalities for ED were outlined in detail; the numbers in parentheses indicate the absolute and relative numbers of studies identified in our systematic review in which ESWT was combined with these treatment modalities: PDE5i (21; 24%), physiotherapeutic exercises (1; 1%), L-arginine supplementation (2; 2%) and vacuum pump rehabilitation (1, 1%). No study combined ESWT with intracavernosal self-injection therapy, medicated urethral system for erection; lifestyle modification, L-citrulline supplementation, ginseng, Vitamin D, curcumin, or psychotherapy/counseling, respectively.

It is beyond the scope of this review to develop a comprehensive strategy for combining ESWT with other treatment modalities in the management of ED. In particular, this would require a thorough search for potential matches between the pathophysiology of certain subtypes of ED (e.g., vasculogenic, post-prostatectomy) with the molecular and cellular mechanisms of action of ESWs and the aforementioned other treatment modalities on connective, muscle, and nerve tissue. However, it must be considered that the mere description of a mechanism of action for ESWT in a study does not necessarily imply that this mechanism is relevant to the observed clinical effects.^13^

Clinical studies on ESWT for ED that do not account for patients’ blood testosterone levels should be considered outdated and should not be included in future evaluations of this treatment modality. Furthermore, ESWT for ED should always be combined with pelvic floor muscle training, which can provide additional benefits by targeting specific areas relevant to erectile dysfunction.^3^ Given that ESWT is highly effective in treating functional/ultrastructural muscle injuries^116^ it is reasonable to hypothesize that, in addition to applying ESWs on penile tissue, targeting the pelvic floor muscles with ESWs (as performed in^47^) may further improve the efficacy of ESWT for ED.

In summary, the possibilities for clinical researchers to combine ESWT with other treatment modalities in the management of ED are virtually limitless. The available literature does not indicate any possible mutual blockages of the mechanisms of action of ESWT and the other treatment modalities mentioned, as is the case, e.g., with the combination of ESWT and certain muscle relaxants in the treatment of chronic, nonspecific low back pain^117^ or the combination of ESWT and diclofenac (a nonsteroidal anti-inflammatory drug) in the treatment of structural muscle injuries.^118^ However, the consistent implementation of the other two aspects of ESWT for ED discussed here (adequate characterization of ESWs, appropriate handling of missing data and intercurrent events) seems to us at present to be more important than considering possible combination therapies.

## LIMITATIONS

The main limitation of this systematic review is that the impact of the substantial heterogeneity of the investigated variables (type of ED, response to PDE5i, IIEF-EF scores at baseline, patient’s age, duration of ED before baseline, type of applied ESWs, details of the treatment protocols) on the treatment outcome in the 87 assessed studies was not further discussed. It is correct that (i) younger patients may respond differently to ESWT for ED than older patients, (ii) ED post-prostatectomy and vasculogenic ED have a different pathophysiology that may require different treatment protocols, and (iii) patients suffering from ED for 20 years may experience different outcome of ESWT than patients suffering from ED for a few months, etc. Conversely, reported differences in treatment outcome may be substantially biased by profound differences in the 3D acoustic pressure fields of the applied ESWs (even if the specified energy density was the same) and inappropriate handling of missing data and intercurrent events. As in case of combination therapies these issues should be addressed first before analyzing the impact of differences in types of ED, subgroups of patients and treatment protocols on the outcome of ESWT for ED.

## CONCLUSION

Current assessments of ESWT for ED as investigational (by, e.g., the AUA; Evidence Level: Grade C) may not stem from a lack of clinical studies, insufficient related basic science, or an inadequate number of systematic reviews and meta-analyses. Instead, the deficits lie in the area of the scientific quality of the clinical studies published to date, as detailed in this systematic review. We hypothesize that this unfortunate situation will only change if the following aspects will be rigorously considered in future clinical studies on ESWT for ED: adequate characterization of ESWs, appropriate handling of missing data and intercurrent events, and comprehensive classification of ESWT in the overall context of the available treatment options for ED. Conversely, we are convinced that the consistent implementation of these aspects will significantly contribute to establishing ESWT as the first truly regenerative therapy in the management of ED. This overall aim justifies the corresponding efforts, for the benefit of our patients.

## Author contributions

Janak Desai (Conceptualization, Writing – review & editing), Eric Huyghe (Conceptualization, Writing – review & editing), Gayle Maffulli (Conceptualization, Writing – review & editing), Carmen Nussbaum-Krammer (Conceptualization, Formal Analysis, Writing – review & editing), Jessica Tittelmeier (Conceptualization, Formal Analysis, Writing – review & editing), Christoph Schmitz (Conceptualization, Data curation, Formal analysis, Methodology, Project administration, Software, Visualization, Writing – original draft).

## Supplementary material

Supplementary material is available at the end of this preprint.

## Data availability

The data underlying this article are available in the article and in its online supplementary material.

## Supplementary Data

This Online Supplement contains the raw data of 55 variables extracted from 87 clinical trials on extracorporeal shock wave therapy for erectile dysfunction published until September 27, 2024.

## Abbreviations used in Supplementary Data

3D: Three-dimensional
BOCF: Baseline observation carried forward
C: Control treatment
D: Day
ED^+^: Positive energy density
ED^total^: Total energy density
EH: Electrohydraulic
EM: Electromagnetic
EM-B: Electromagnetic-ballistic
ESW: Extracorporeal shock wave
ESWT: Extracorporeal shock wave therapy
F: Focused
Hz: Hertz
IIEF-EF: International Index of Erectile Function - Erectile Domain
ITT: Intent-to-treat
KL: Kilovolt
L: Linear
mJ/mm^2^: Millijoule per squared millimeter
n.a.: Not applicable
n.s.: Not specified
P-B: Pneumatic-ballistic
PDE5i: Phosphodiesterase-5 inhibitors
PE: Piezoelectric
PRP: Platelet derived plasma
R: Radial
Ref: Reference
RCT: Randomized controlled trial
SD: Standard deviation
TMS: Transcranial magnetic stimulation
U: Unfocused
V: Variable
W: Week

---

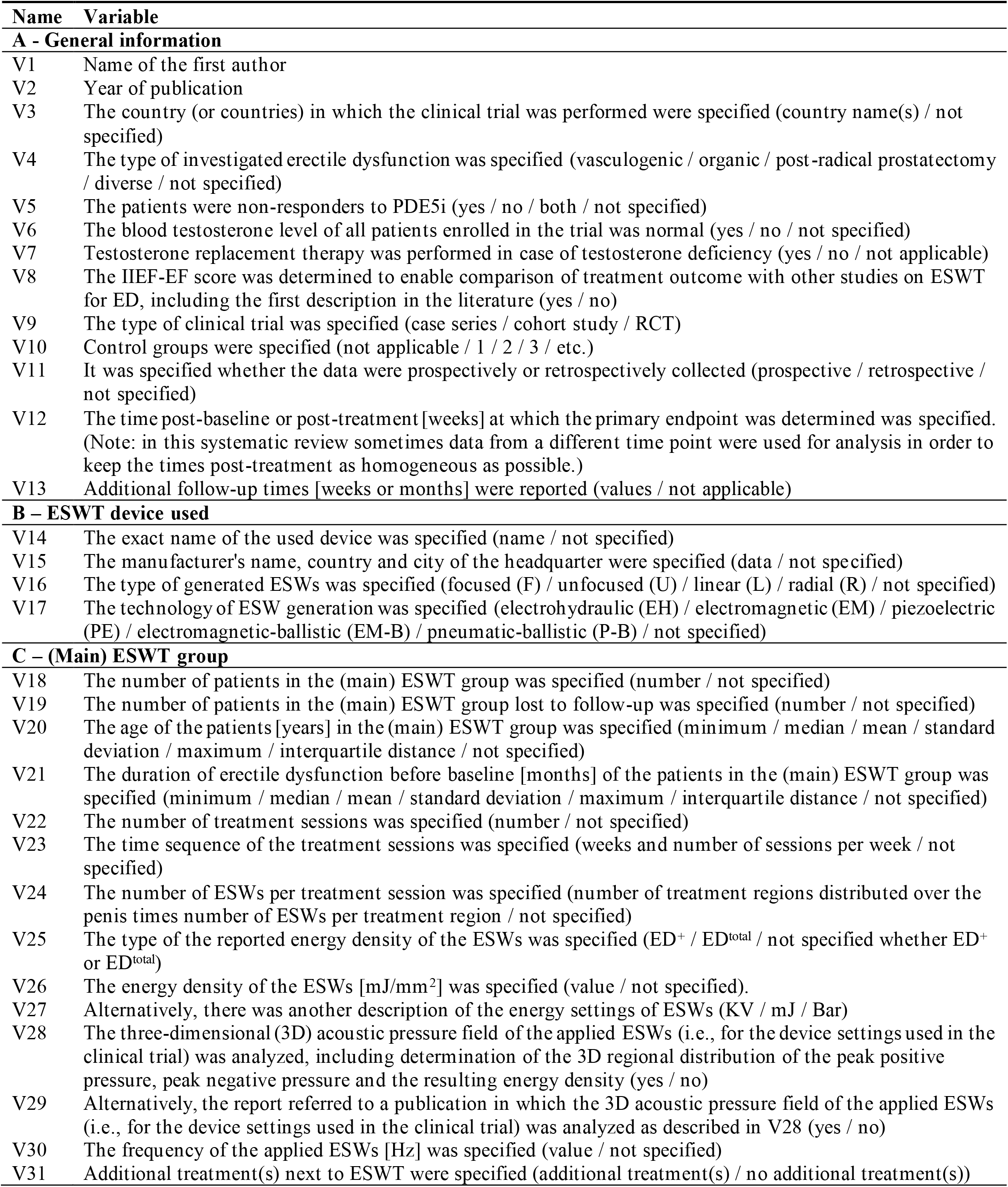
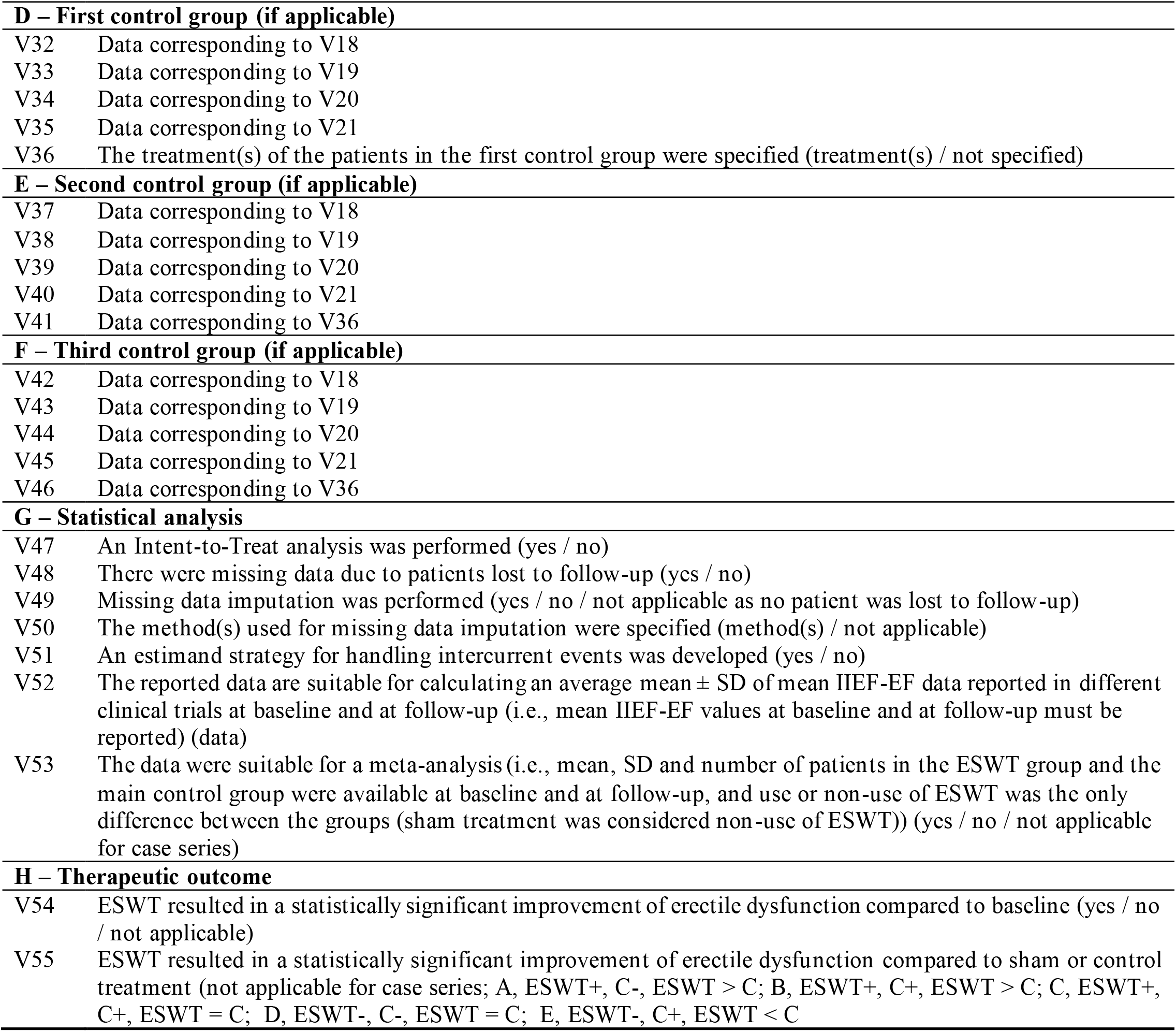

| Name | Variable |
| --- | --- |
| **A - General information** |  |
| V1 | Name of the first author |
| V2 | Year of publication |
| V3 | The country (or countries) in which the clinical trial was performed were specified (country name(s) / not specified) |
| V4 | The type of investigated erectile dysfunction was specified (vasculogenic / organic / post-radical prostatectomy / diverse / not specified) |
| V5 | The patients were non-responders to PDE5i (yes / no / both / not specified) |
| V6 | The blood testosterone level of all patients enrolled in the trial was normal (yes / no / not specified) |
| V7 | Testosterone replacement therapy was performed in case of testosterone deficiency (yes / no / not applicable) |
| V8 | The IIEF-EF score was determined to enable comparison of treatment outcome with other studies on ESWT for ED, including the first description in the literature (yes / no) |
| V9 | The type of clinical trial was specified (case series / cohort study / RCT) |
| V10 | Control groups were specified (not applicable / 1 / 2 / 3 / etc.) |
| V11 | It was specified whether the data were prospectively or retrospectively collected (prospective / retrospective / not specified) |
| V12 | The time post-baseline or post-treatment [weeks] at which the primary endpoint was determined was specified. (Note: in this systematic review sometimes data from a different time point were used for analysis in order to keep the times post-treatment as homogeneous as possible.) |
| V13 | Additional follow-up times [weeks or months] were reported (values / not applicable) |
| **B – ESWT device used** |  |
| V14 | The exact name of the used device was specified (name / not specified) |
| V15 | The manufacturer's name, country and city of the headquarter were specified (data / not specified) |
| V16 | The type of generated ESWs was specified (focused (F) / unfocused (U) / linear (L) / radial (R) / not specified) |
| V17 | The technology of ESW generation was specified (electrohydraulic (EH) / electromagnetic (EM) / piezoelectric (PE) / electromagnetic-ballistic (EM-B) / pneumatic-ballistic (P-B) / not specified) |
| **C – (Main) ESWT group** |  |
| V18 | The number of patients in the (main) ESWT group was specified (number / not specified) |
| V19 | The number of patients in the (main) ESWT group lost to follow-up was specified (number / not specified) |
| V20 | The age of the patients [years] in the (main) ESWT group was specified (minimum / median / mean / standard deviation / maximum / interquartile distance / not specified) |
| V21 | The duration of erectile dysfunction before baseline [months] of the patients in the (main) ESWT group was specified (minimum / median / mean / standard deviation / maximum / interquartile distance / not specified) |
| V22 | The number of treatment sessions was specified (number / not specified) |
| V23 | The time sequence of the treatment sessions was specified (weeks and number of sessions per week / not specified) |
| V24 | The number of ESWs per treatment session was specified (number of treatment regions distributed over the penis times number of ESWs per treatment region / not specified) |
| V25 | The type of the reported energy density of the ESWs was specified ($ED^+$ / $ED^{total}$ / not specified whether $ED^+$ or $ED^{total}$) |
| V26 | The energy density of the ESWs [$mJ/mm^2$] was specified (value / not specified). |
| V27 | Alternatively, there was another description of the energy settings of ESWs (KV / mJ / Bar) |
| V28 | The three-dimensional (3D) acoustic pressure field of the applied ESWs (i.e., for the device settings used in the clinical trial) was analyzed, including determination of the 3D regional distribution of the peak positive pressure, peak negative pressure and the resulting energy density (yes / no) |
| V29 | Alternatively, the report referred to a publication in which the 3D acoustic pressure field of the applied ESWs (i.e., for the device settings used in the clinical trial) was analyzed as described in V28 (yes / no) |
| V30 | The frequency of the applied ESWs [Hz] was specified (value / not specified) |
| V31 | Additional treatment(s) next to ESWT were specified (additional treatment(s) / no additional treatment(s)) |

| **Name** | **Variable** |
| --- | --- |
| **D – First control group (if applicable)** |  |
| V32 | Data corresponding to V18 |
| V33 | Data corresponding to V19 |
| V34 | Data corresponding to V20 |
| V35 | Data corresponding to V21 |
| V36 | The treatment(s) of the patients in the first control group were specified (treatment(s) / not specified) |
| **E – Second control group (if applicable)** |  |
| V37 | Data corresponding to V18 |
| V38 | Data corresponding to V19 |
| V39 | Data corresponding to V20 |
| V40 | Data corresponding to V21 |
| V41 | Data corresponding to V36 |
| **F – Third control group (if applicable)** |  |
| V42 | Data corresponding to V18 |
| V43 | Data corresponding to V19 |
| V44 | Data corresponding to V20 |
| V45 | Data corresponding to V21 |
| V46 | Data corresponding to V36 |
| **G – Statistical analysis** |  |
| V47 | An Intent-to-Treat analysis was performed (yes / no) |
| V48 | There were missing data due to patients lost to follow-up (yes / no) |
| V49 | Missing data imputation was performed (yes / no / not applicable as no patient was lost to follow-up) |
| V50 | The method(s) used for missing data imputation were specified (method(s) / not applicable) |
| V51 | An estimand strategy for handling intercurrent events was developed (yes / no) |
| V52 | The reported data are suitable for calculating an average mean ± SD of mean IIEF-EF data reported in different clinical trials at baseline and at follow-up (i.e., mean IIEF-EF values at baseline and at follow-up must be reported) (data) |
| V53 | The data were suitable for a meta-analysis (i.e., mean, SD and number of patients in the ESWT group and the main control group were available at baseline and at follow-up, and use or non-use of ESWT was the only difference between the groups (sham treatment was considered non-use of ESWT)) (yes / no / not applicable for case series) |
| **H – Therapeutic outcome** |  |
| V54 | ESWT resulted in a statistically significant improvement of erectile dysfunction compared to baseline (yes / no / not applicable) |
| V55 | ESWT resulted in a statistically significant improvement of erectile dysfunction compared to sham or control treatment (not applicable for case series; A, ESWT+, C-, ESWT > C; B, ESWT+, C+, ESWT > C; C, ESWT+, C+, ESWT = C; D, ESWT-, C-, ESWT = C; E, ESWT-, C+, ESWT < C) |

| Ref | V1 | V2 | V3 | V4 | V5 | V6 | V7 | V8 |
| --- | --- | --- | --- | --- | --- | --- | --- | --- |
| 1 | İbis | 2024 | Turkey | Not specified | Yes | Yes | Not applicable | No |
| 2 | Kalyvianakis | 2024 | Greece | Vasculogenic | No | Yes | Not applicable | Yes |
| 3 | Kaynak | 2024 | Turkey | Organic | Both | Not tested | Not considered | No |
| 4 | Rubino | 2024 | Italy | Vasculogenic | No | Yes | Not applicable | No |
| 5 | Trishch | 2024 | Ukraine | Not specified | No | Not tested | Not considered | Yes |
| 6 | Bayraktar | 2023 | Turkey | Not specified | Yes | Yes | Not applicable | No |
| 7 | Bryk | 2023 | USA | P-RP | No | Not tested | Not considered | No |
| 8 | Islam | 2023 | Bangladesh | Not specified | No | Not tested | Not considered | No |
| 9 | Jang | 2023 | Korea | P-RP | No | Not tested | Not considered | No |
| 10 | Kennady | 2023 | USA | Organic | Both | Yes | Not applicable | No |
| 11 | Kohada | 2023 | Japan | P-RP | No | Not tested | Not considered | Yes |
| 12 | Kurosawa | 2023 | Japan | Diverse | No | Yes | Not applicable | No |
| 13 | Ma | 2023 | China | Organic | No | Not tested | Not considered | No |
| 14 | Vena | 2023 | Italy | Organic | Yes | Yes | Not applicable | Yes |
| 15 | Wang | 2023 | China | Diverse | No | Not tested | Not considered | Yes |
| 16 | Cocci | 2022 | Italy | <sup>1</sup> | No | Yes | Not applicable | No |
| 17 | Daneshwar | 2022 | Malaysia | <sup>2</sup> | No | Not tested | Not considered | No |
| 18 | Ergün | 2022 | Turkey | Vasculogenic | Yes | Not tested | Not considered | No |
| 19 | Gallo | 2022 | Italy | Not specified | No | Yes | Not applicable | Yes |
| 20 | Geyik | 2022 | Turkey | Not specified | No | Yes | Not applicable | Yes |
| 21 | Kalyvianakis | 2022 | Greece | Vasculogenic | No | Yes | Not applicable | Yes |
| 22 | Motil | 2022 | Czech republic | P-RP | No | Yes | Not applicable | No |
| 23 | Mykoniatis | 2022 | Greece | Vasculogenic | No | Yes | Not applicable | Yes |
| 24 | Oginski | 2022 | Germany | Organic | Yes | Not tested | Not considered | No |
| 25 | Ong | 2022 | Malaysia | Diverse | No | Not tested | Not considered | No |
| 26 | Sandoval-Salinas | 2022 | Colombia | Organic | No | Not tested | Not considered | Yes |
| 27 | Scroppa | 2022 | Italy | Vasculogenic | Yes | Yes | Not applicable | No |
| 28 | Tao | 2022 | China | Vasculogenic | Yes | Yes | Not applicable | Yes |
| 29 | Zanaty | 2022 | Egypt | Organic | No | Yes | Not applicable | No |
| 30 | Adeldaeim | 2021 | Egypt | Vasculogenic | No | No | Yes | No |
| 31 | Akande | 2021 | Nigeria | Vasculogenic | No | Not tested | Not considered | No |
| 32 | Caretta | 2021 | Italy | Organic | No | Yes | Not applicable | No |
| 33 | Chung (follow-up) | 2021 | Australia | Vasculogenic | No | Not tested | Not considered | No |
| 34 | Geyik | 2021 | Turkey | Diverse | Yes | Yes | Not applicable | Yes |
| 35 | Karakose | 2021 | Turkey | P-RP | No | Not tested | Not considered | No |
| 36 | Ladegaard | 2021 | Denmark | P-RP | No | Not tested | Not considered | No |
| 37 | Lau | 2021 | Singapore | Diverse | No | Yes | Not applicable | No |
| 38 | Lei | 2021 | China | Diverse | No | Not tested | Not considered | No |
| 39 | Ortac | 2021 | Turkey | Vasculogenic | No | Yes | Not applicable | Yes |
| 40 | Palmieri | 2021 | Italy | Vasculogenic | Yes | Not tested | Not considered | Yes |
| 41 | Shendy | 2021 | Egypt | Vasculogenic | No | Not tested | Not considered | Yes |
| 42 | Tzou | 2021 | Taiwan | Diverse | Yes | Yes | Not applicable | No |
| 43 | Vinay | 2021 | Spain | Diverse | Yes | Not tested | Not considered | Yes |
| 44 | Baccaglini | 2020 | Brazil | P-RP | No | Not tested | Not considered | No |
| 45 | Eryilmaz | 2020 | Turkey | Vasculogenic | Yes | Yes | Not applicable | No |
| 46 | Huang | 2020 | China | Diverse | No | Yes | Not applicable | No |
| 47 | Kalyvianakis | 2020 | Greece | Vasculogenic | No | Not tested | Not considered | Yes |
| 48 | Konchugova | 2020 | Russia | Vasculogenic | No | Not tested | Not considered | No |
| 49 | Lurz | 2020 | USA | Vasculogenic | No | Yes | Not applicable | Yes |
| 50 | Inoue | 2020 | Japan | P-RP | No | Yes | Not applicable | No |

<sup>1</sup> Priapism-induced ED (45 days post-emergency management)
<sup>2</sup> ED on the basis of chronic pelvic pain syndrome

| Ref | V1 | V2 | V3 | V4 | V5 | V6 | V7 | V8 |
| --- | --- | --- | --- | --- | --- | --- | --- | --- |
| 51 | Kim | 2020 | Korea | Not specified | No | Not tested | Not considered | Yes |
| 52 | Patel | 2020 | USA | Organic | No | Yes | Not applicable | Yes |
| 53 | Verze | 2020 | Italy | Vasculogenic | No | Not tested | Not considered | No |
| 54 | Wang | 2020 | China | Organic | Both | Yes | Not applicable | Yes |
| 55 | Wu | 2020 | USA | Vasculogenic | No | Not tested | Not considered | No |
| 56 | Zasieda | 2020 | Ukraine | <sup>3</sup> | No | Yes | Not applicable | No |
| 57 | Costa | 2019 | Portugal | Organic | No | Not tested | Not considered | No |
| 58 | Musa | 2019 | Egypt | Vasculogenic | Yes | No | Yes | <sup>4</sup> |
| 59 | Sramkova | 2019 | Czech republic | Vasculogenic | No | Yes | Not applicable | No |
| 60 | Vita | 2019 | Italy | Vasculogenic | Yes | Yes | Not applicable | No |
| 61 | Wang | 2019 | Taiwan | <sup>5</sup> | Yes | Not tested | Not considered | No |
| 62 | Yamaçake | 2019 | Brazil | <sup>6</sup> | No | Yes | Not applicable | No |
| 63 | De Oliveira | 2018 | Portugal | Diverse | No | Not tested | Not considered | No |
| 64 | Fojecki | 2018 | Demnark | Organic | No | Yes | Not applicable | Yes |
| 65 | Kalyvianakis | 2018 | Greece | Vasculogenic | No | Not tested | Not considered | Yes |
| 66 | Kitrey | 2018 | Israel | Diverse | No | Not tested | Not considered | <sup>4</sup> |
| 67 | Zewin | 2018 | Egypt | P-RP | No | Not tested | Not considered | No |
| 68 | Ayala | 2017 | Colombia / Spain | Vasculogenic | No | Not tested | Not considered | No |
| 69 | Fojecki | 2017 | Denmark | Organic | No | Yes | Not applicable | Yes |
| 70 | Kalyvianakis | 2017 | Greece | Vasculogenic | No | Not tested | Not considered | Yes |
| 71 | Tsai | 2017 | Taiwan | Organic | Yes | No | Yes | No |
| 72 | Bechara | 2016 | Argentina | Vasculogenic | Yes | Not tested | Not considered | <sup>7</sup> |
| 73 | Frey | 2016 | Denmark | P-RP | No | Not tested | Not considered | No |
| 74 | Hisasue | 2016 | Japan | Diverse | No | Yes | Not applicable | No |
| 75 | Kitrey | 2016 | Israel | Vasculogenic | Yes | Not tested | Not considered | Yes |
| 76 | Bechara | 2015 | Argentina | Vasculogenic | Yes | Not tested | Not considered | Yes |
| 77 | Chung | 2015 | Australia | Vasculogenic | No | Not tested | Not considered | No |
| 78 | Olsen | 2015 | Denmark | Organic | No | Not tested | Not considered | No |
| 79 | Pelayo-Nieto | 2015 | Mexico | Vasculogenic | No | Not tested | Not considered | Yes |
| 80 | Reisman | 2015 | <sup>8</sup> | Vasculogenic | No | Not tested | Not considered | Yes |
| 81 | Ruffo | 2015 | Italy | Organic | Yes | Not tested | Not considered | Yes |
| 82 | Srini | 2015 | India | Organic | No | Not tested | Not considered | Yes |
| 83 | Yee | 2014 | China | Organic | No | Not tested | Not considered | Yes |
| 84 | Gruenwald | 2012 | Israel | Vasculogenic | No | Not tested | Not considered | Yes |
| 85 | Palmieri | 2012 | Italy | ED + PD | No | Not tested | Not considered | No |
| 86 | Vardi | 2012 | Israel | Vasculogenic | No | Not tested | Not considered | Yes |
| 87 | Vardi | 2010 | Israel | Vasculogenic | No | Not tested | Not considered | Yes |

<sup>3</sup> Veno-occlusive ED based on hypogonadotropic hypogonadism
<sup>4</sup> No IIEF-EF score at any follow-up time (M1, M6, M12, M18) reported
<sup>5</sup> post pelvic fractures associated with urethral injury
<sup>6</sup> ED in kidney transplant recipients (multifactorial)
<sup>7</sup> Only data of "LI-ESWT responders" reported (24 of 40 analyzed patients, with 50 patients included in the study and 10 patients lost to follow-up)
<sup>8</sup> Netherlands, Palestine, Lithuania and Czech Republic

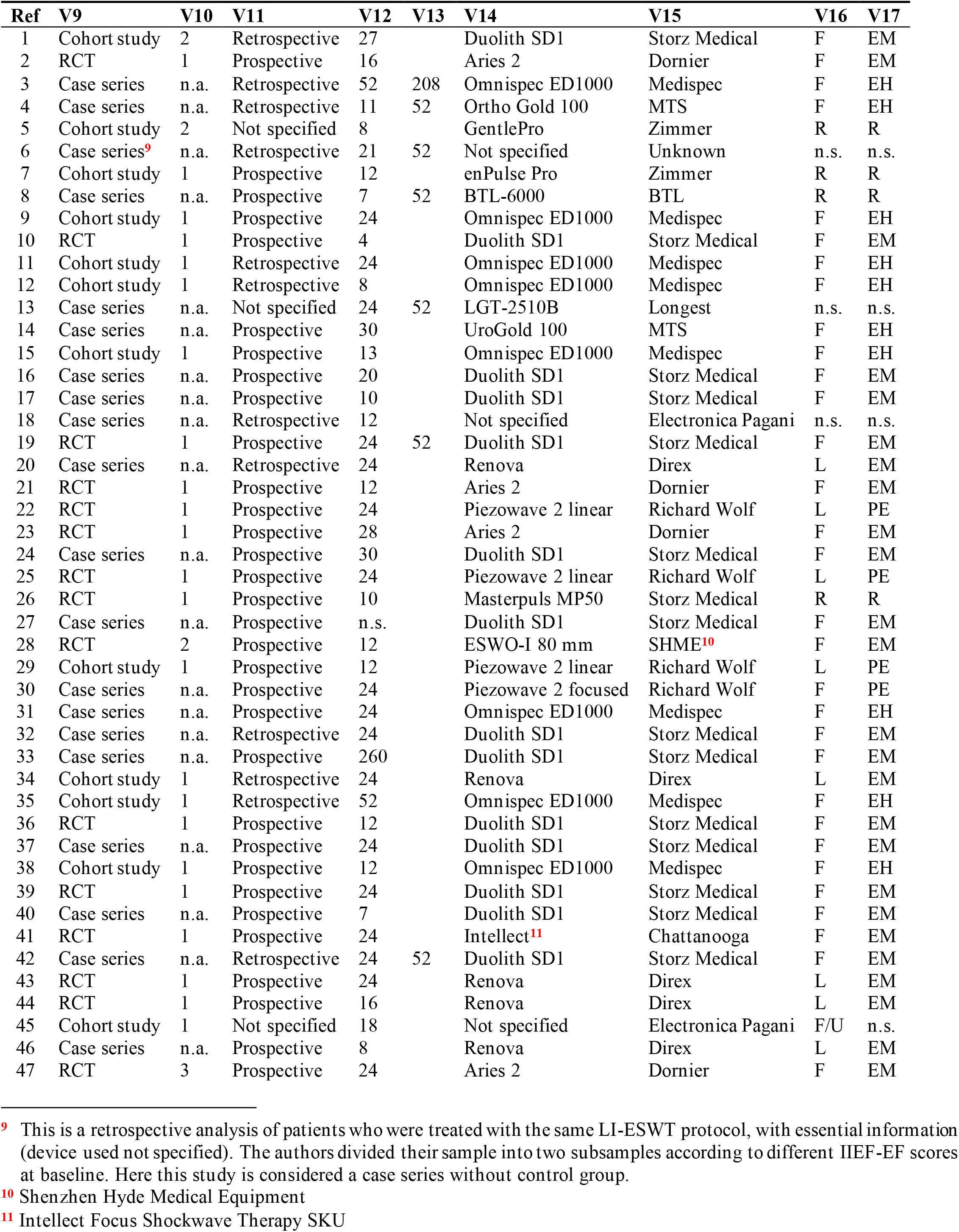
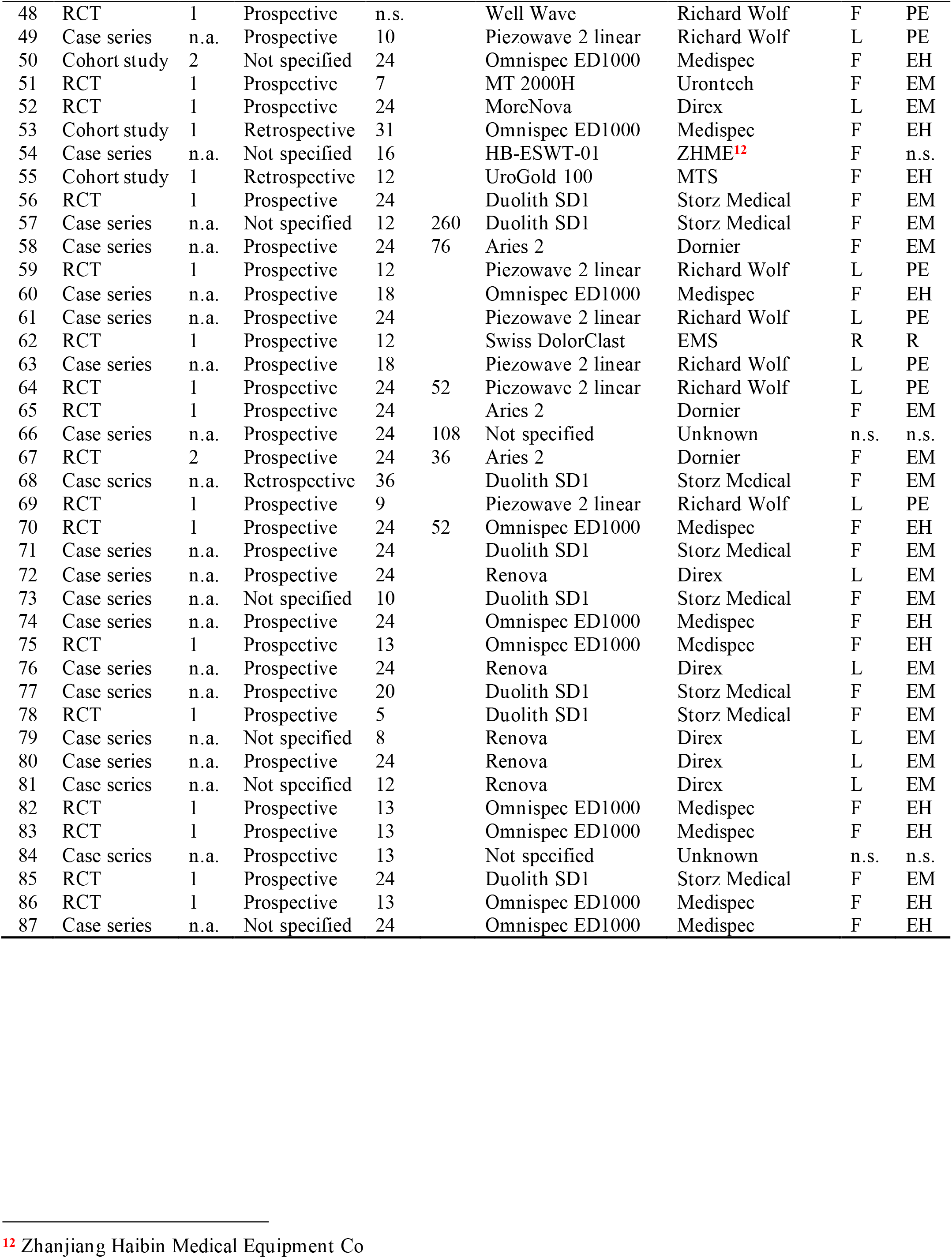

| Ref | V18 | V19 | V20<sup>13</sup> | V20<sup>14</sup> | V20<sup>15</sup> | V20<sup>16</sup> | V21<sup>13</sup> | V21<sup>14</sup> | V21<sup>15</sup> | V21<sup>17</sup> |
| --- | --- | --- | --- | --- | --- | --- | --- | --- | --- | --- |
| 1 | 36 | 0 | 56.1 |  | 7.3 |  |  |  |  | >6 |
| 2 | 34 | 0 | 59 |  | 8.6 |  | 94 |  | 75 |  |
| 3 | 116 | 0 | 47.3 |  |  | 19-71 |  |  |  | <6 |
| 4 | 113 | 0 |  | 50 |  |  | n.s. |  |  |  |
| 5 | 34 | 0 | 35.3 |  |  | 25-50 | 23.4 |  |  | 6-84 |
| 6 | 126 | 0 | 50.5 |  | 12.4 |  |  |  |  | >6 |
| 7 | 44 | 1 | 59.5 |  | 6.9 |  | n.a. |  |  |  |
| 8 | 31 | 0 | 44.6 |  |  | 25-78 |  |  |  | >6 |
| 9 | 56 | 15 | 62 |  |  |  | n.a. |  |  |  |
| 10 | 17 | 0 | 67 |  |  |  | n.s. |  |  |  |
| 11 | 16 | <sup>18</sup> | 63 |  |  | 52-72 | n.a. |  |  |  |
| 12 | 76 | 0 | 52.8 |  | 11.7 |  | n.s. |  |  |  |
| 13 | 26 | 0 | 43.8 |  | 8.1 |  | n.s. |  |  |  |
| 14 | 21 | 3 | 56.3 |  | 9.1 |  | >12 |  |  |  |
| 15 | 51 | 9 | 33.9 |  | 6.2 |  | 24 |  |  | 12-36 |
| 16 | 5 | 0 | 31.2 |  |  | 24-36 | n.s. |  |  |  |
| 17 | 50 | 0 | 41.9 |  | 11.7 |  | n.s. |  |  |  |
| 18 | 63 | 0 | 57.1 |  | 5.1 |  | 25 |  | 5.5 |  |
| 19 | 50 | 9 | 50.5 |  | 13.7 |  |  |  |  | >3 |
| 20 | 41 | 0 | 51.6 |  | 11.8 |  | 54.5 |  | 33 |  |
| 21 | 35 | 1 |  | 54 |  |  |  | 68 |  |  |
| 22 | 20 | 4 | 58.3 |  | 4.9 |  |  | 7 |  | 3-18 |
| 23 | 25 | 0 |  | 56 |  |  |  | 44 |  |  |
| 24 | 50 | 0 |  | 59 |  |  |  | 48 |  |  |
| 25 | 27 | 0 |  | 61 |  |  |  | 24 |  |  |
| 26 | 40 | 3 | 47.7 |  | 12.8 |  | 5.5 |  | 6.6 |  |
| 27 | 30 | 0 | 52.8 |  | 7.9 |  | n.s. |  |  |  |
| 28 | 35 | 2 | 48.3 |  | 3.5 |  | 45.3 |  | 25.1 |  |
| 29 | 25 | 0 | 43.8 |  |  | 28-62 | 66 |  |  | 12-180 |
| 30 | 425 | 0 |  | 55 |  | 30-70 | 25 |  |  | 12-38 |
| 31 | 30 | 8 | 53.7 |  | 8 |  |  |  |  | >6 |
| 32 | 111 | 0 | 53.7 |  | 11.6 |  | n.s. |  |  |  |
| 33 | 30 | 0 | 55.8 |  |  | 42-68 | 21.8 |  |  | 6-60 |
| 34 | 91 | 0 | 46.9 |  | 11.9 |  | 48 |  | 32 |  |
| 35 | 34 | 0 | 59.2 |  |  | 51-70 | n.a. |  | -- |  |
| 36 | 20 | 0 | 60.8 |  | 6.8 |  | 11.2 |  | 4.5 |  |
| 37 | 19 | 8 | 57.4 |  |  | 38-70 | 46.3 |  |  | 12-120 |
| 38 | 60 | 14 | 33.4 |  | 6.2 |  | 24 |  |  | 12-36 |
| 39 | 44 | 25 | 42.3 |  | 9.9 |  | 33.7 |  | 36.5 |  |
| 40 | 109 | 3 | 57.9 |  |  | 21-78 | 33.3 |  |  | 3-103 |
| 41 | 21 | 0 | 48.6 |  | 5.5 |  | 118 |  | 13 |  |
| 42 | 69 | 0 |  | 55 |  | 45-66 |  | 12 |  | 6-36 |
| 43 | 40 | 0 |  | 60 |  | 54-66 |  | 36 |  | 24-72 |
| 44 | 46 | 10 | 64.6 |  | 5.3 |  | n.a. |  |  |  |
| 45 | 20 | 0 | 45.6 |  |  |  | n.s. |  |  |  |

<sup>13</sup> Mean
<sup>14</sup> Median
<sup>15</sup> Standard deviation
<sup>16</sup> Range
<sup>17</sup> Minimum value or range

| Ref | V18 | V19 | V20<sup>13</sup> | V20<sup>14</sup> | V20<sup>15</sup> | V20<sup>16</sup> | V21<sup>13</sup> | V21<sup>14</sup> | V21<sup>15</sup> | V21<sup>17</sup> |
| --- | --- | --- | --- | --- | --- | --- | --- | --- | --- | --- |
| 46 | 35 | 0 | 33 |  |  | 21-68 |  |  |  | >6 |
| 47 | 24 | 1 | 58.7 |  | 9.5 |  |  |  |  | >6 |
| 48 | 20 | 0 | 59.2 |  | 8.7 |  | n.s. |  |  |  |
| 49 | 25 | 3 |  | 60 |  | 55-69 |  | 60 |  | 18-222 |
| 50 | 5 | <sup>19</sup> | 62.2 |  | 2.7 |  | n.a. |  |  |  |
| 51 | 46 | 8 | 63.2 |  | 5.4 |  |  |  |  | >6 |
| 52 | 45 | 5 | 53.2 |  |  | 30-78 |  |  |  | >6 |
| 53 | 78 | 0 | 56 |  | 9.6 |  | n.s. |  |  |  |
| 54 | 45 | 0 |  | 60 |  | 30-84 |  | 38 |  | 6-120 |
| 55 | 24 | 0 |  | 61 |  | 48-70 |  | 61 |  | 36-61 |
| 56 | 22 | 0 | 51 |  |  | 45-60 | n.s. |  |  |  |
| 57 | 18 | 0 | 61.1 |  | 7.2 |  |  |  |  | >12 |
| 58 | 55 | 3 | 51 |  |  | 23-74 | 34.5 |  |  | 9-60 |
| 59 | 30 | 0 | 53.9 |  | 9.3 |  | 42 |  |  | 6-204 |
| 60 | 20 | 0 | 58.5 |  | 10.3 |  | 98.4 |  | 80.4 |  |
| 61 | 12 | 0 | 35.5 |  | 7.1 |  |  |  |  | >6 |
| 62 | 10 | 0 | 55.1 |  |  | 47-60 | 22.7 |  | 9.2 |  |
| 63 | 25 | 0 |  | 61 |  | 27-73 | 24 |  |  |  |
| 64 | 63 | 11 | 65.4 |  | 7.9 |  |  |  |  | >6 |
| 65 | 21 | 3 | 57.5 |  | 10 |  |  |  |  | >3 |
| 66 | 156 | 0 | 59.1 |  | 10.1 |  | 64.6 |  | 49 |  |
| 67 | 49 | 7 | 52.9 |  | 7.2 |  | n.a. |  |  |  |
| 68 | 710 | 298 |  | 58 |  | 24-83 |  |  |  | >3 |
| 69 | 63 | 3 |  | 65.4 | 7.9 |  |  |  |  | >6 |
| 70 | 30 | 0 |  | 53 |  | 31-72 |  | 66 |  | 12-240 |
| 71 | 52 | 0 | 60.1 |  | 11.5 |  | 38.3 |  | 26.9 |  |
| 72 | 25 | 5 | 64.8 |  |  | 48-82 | 70.5 |  | n.s. |  |
| 73 | 18 | 3 |  | 62 |  | 51-70 |  | 24 |  | 12-54 |
| 74 | 56 | 0 |  | 64 |  | 27-83 |  | 36 |  | 6-216 |
| 75 | 40 | 3 |  | 60 |  | 28-27 |  | 60 |  | 11-240 |
| 76 | 25 | 0 |  | 63 |  | 46-78 |  | 42 |  | 12-132 |
| 77 | 30 | 0 | 55.8 |  |  | 42-68 | 21.8 |  |  | 6-60 |
| 78 | 51 | 15 |  | 59 |  | 41-80 |  | 57 |  | 9-240 |
| 79 | 15 | 0 |  | 59.6 |  | 45-70 | n.s. |  |  |  |
| 80 | 58 | 0 | 56.8 |  |  | 33-84 |  |  |  | >3 |
| 81 | 31 | 0 | 59.9 |  | 12.2 |  |  |  |  | >6 |
| 82 | 95 | 35 | n.s. |  |  |  |  |  |  | >6 |
| 83 | 36 | 6 | 58.9 |  | 7.6 |  | 78 |  | 34 |  |
| 84 | 33 | 4 | 61.3 |  |  | 41-79 |  | 60 | n.s. |  |
| 85 | 50 | 0 | 55.5 |  |  | 32-75 | 8.8 |  |  | 6-12 |
| 86 | 46 | 6 |  | 58 |  | 27-72 |  | 42 |  | 6-240 |
| 87 | 20 | 0 | 56.1 |  |  | 33-73 | 34.7 |  | n.s. |  |

<sup>19</sup> Approximately 40% of the patients were lost to follow-up. However, group-specific patients lost to follow-up were not reported.

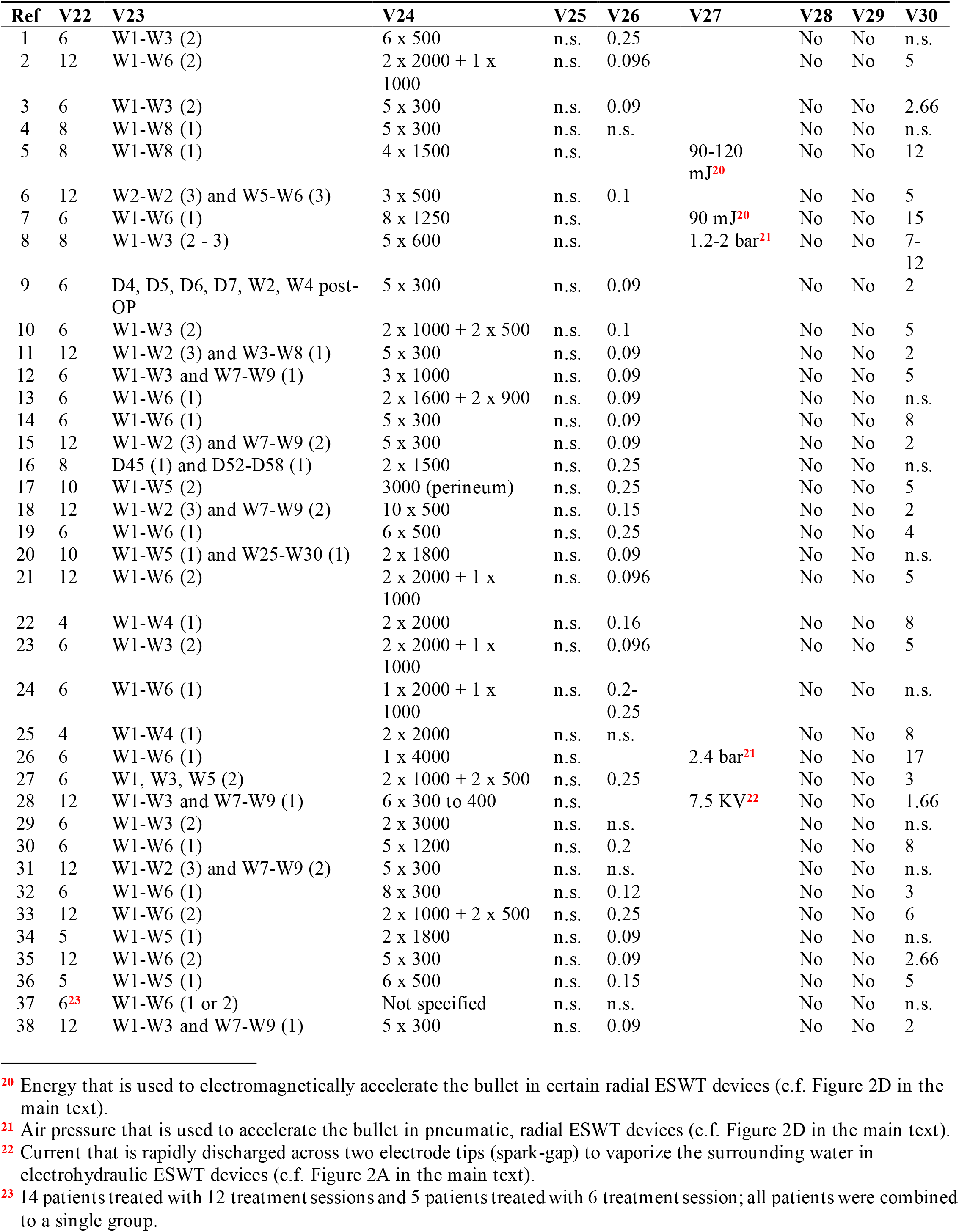
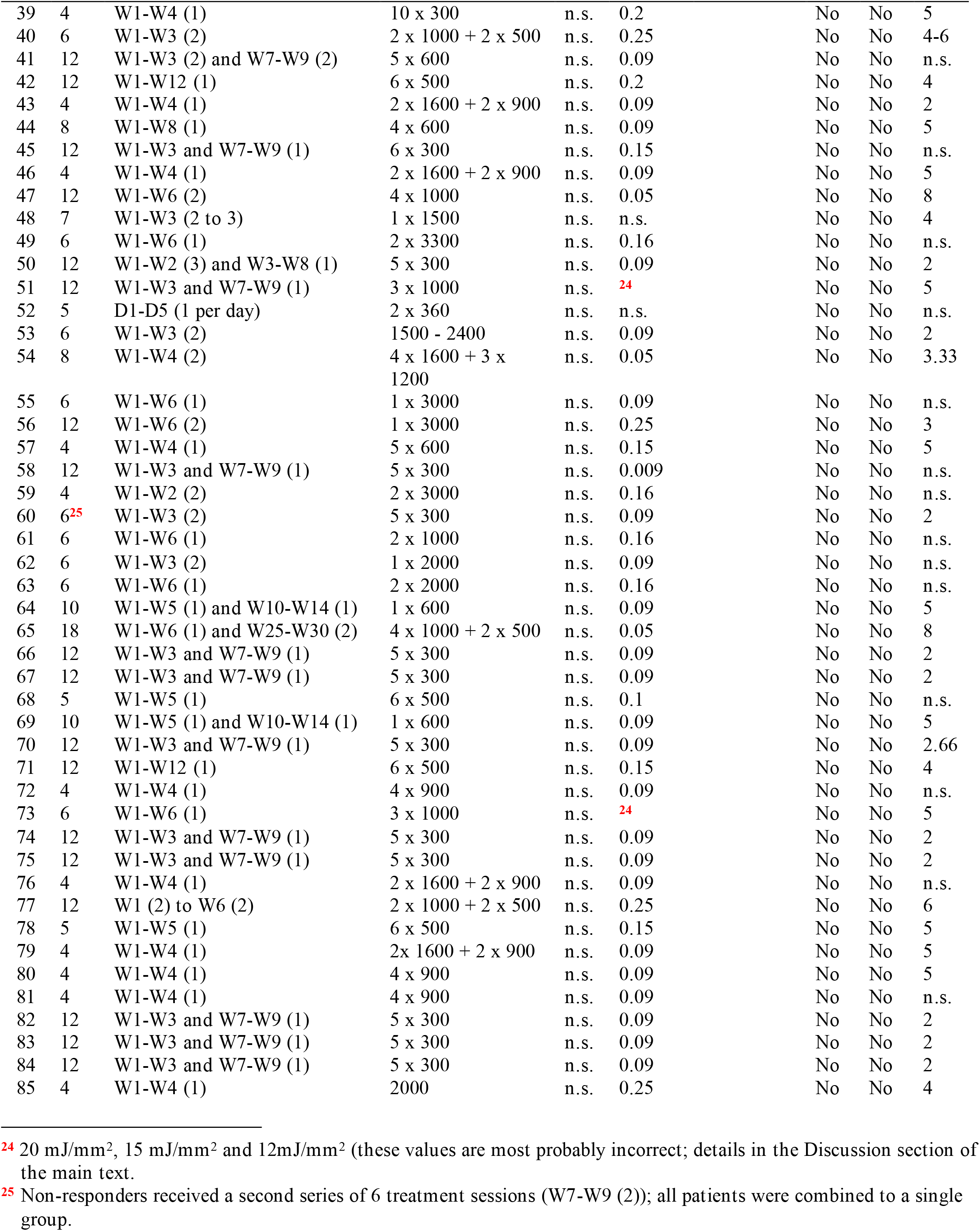
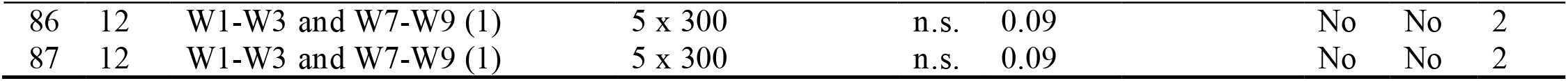

| Ref | V31 |
| --- | --- |
| 1 | PDE5i (20 mg of tadalafil every second day) |
| 2 | PDE5i (daily from W1-W4) |
| 3 | PDE5i (daily for 6 months) |
| 4 | -- |
| 5 | PDE5i + L-arginine |
| 6 | PDE5i (daily for 2 months) |
| 7 | -- |
| 8 | -- |
| 9 | PDE5i (daily for 6 months post-OP) |
| 10 | -- |
| 11 | -- |
| 12 | -- |
| 13 | -- |
| 14 | PDE5i (daily starting at W11) |
| 15 | --- |
| 16 | -- |
| 17 | -- |
| 18 | PDE5i (daily) |
| 19 | PDE5i + L-arginine |
| 20 | -- |
| 21 | -- |
| 22 | -- |
| 23 | PDE5i (daily from W1-W4) |
| 24 | -- |
| 25 | -- |
| 26 | 2000 shots into bilateral ischiocavernosus and bulbospongiosus muscles |
| 27 | -- |
| 28 | Vacuum erectile device (3x /week; 9 weeks) |
| 29 | -- |
| 30 | -- |
| 31 | -- |
| 32 | -- |
| 33 | -- |
| 34 | PRP (3x, W1, W3, W5) |
| 35 | PDE5i (daily) |
| 36 | -- |
| 37 | -- |
| 38 | -- |
| 39 | -- |
| 40 | -- |
| 41 | Kegel exercises three times daily for 6 weeks |
| 42 | -- |
| 43 | -- |
| 44 | PDE5i (daily for 8 weeks post-OP) |
| 45 | -- |
| 46 | -- |
| 47 | -- |
| 48 | TMS |
| 49 | -- |
| 50 | -- |
| 51 | -- |
| 52 | -- |
| 53 | PDE5i (daily for 12 weeks) |
| 54 | PDE5i (daily or on demand) |
| 55 | PDE5i (on demand) |
| 56 | Chorionic gonadotropin + extract of Epimedium Breviconum |
| 57 | -- |
| 58 | -- |
| 59 | -- |
| 60 | PDE5i (daily or on demand) |
| 61 | PDE5i (daily for 6 months post-accident) |
| 62 | -- |
| 63 | PDE5i (daily for 6 weeks) |
| 64 | -- |
| 65 | -- |
| 66 | -- |
| 67 | -- |
| 68 | -- |
| 69 | -- |
| 70 | -- |
| 71 | PDE5i (daily for 12 weeks or on demand) |
| 72 | PDE5i (daily for 4 weeks) |
| 73 | -- |
| 74 | -- |
| 75 | --- |
| 76 | -- |
| 77 | -- |
| 78 | -- |
| 79 | -- |
| 80 | -- |
| 81 | -- |
| 82 | -- |
| 83 | -- |
| 84 | -- |
| 85 | PDE5i (daily for 4 weeks) |
| 86 | -- |
| 87 | -- |

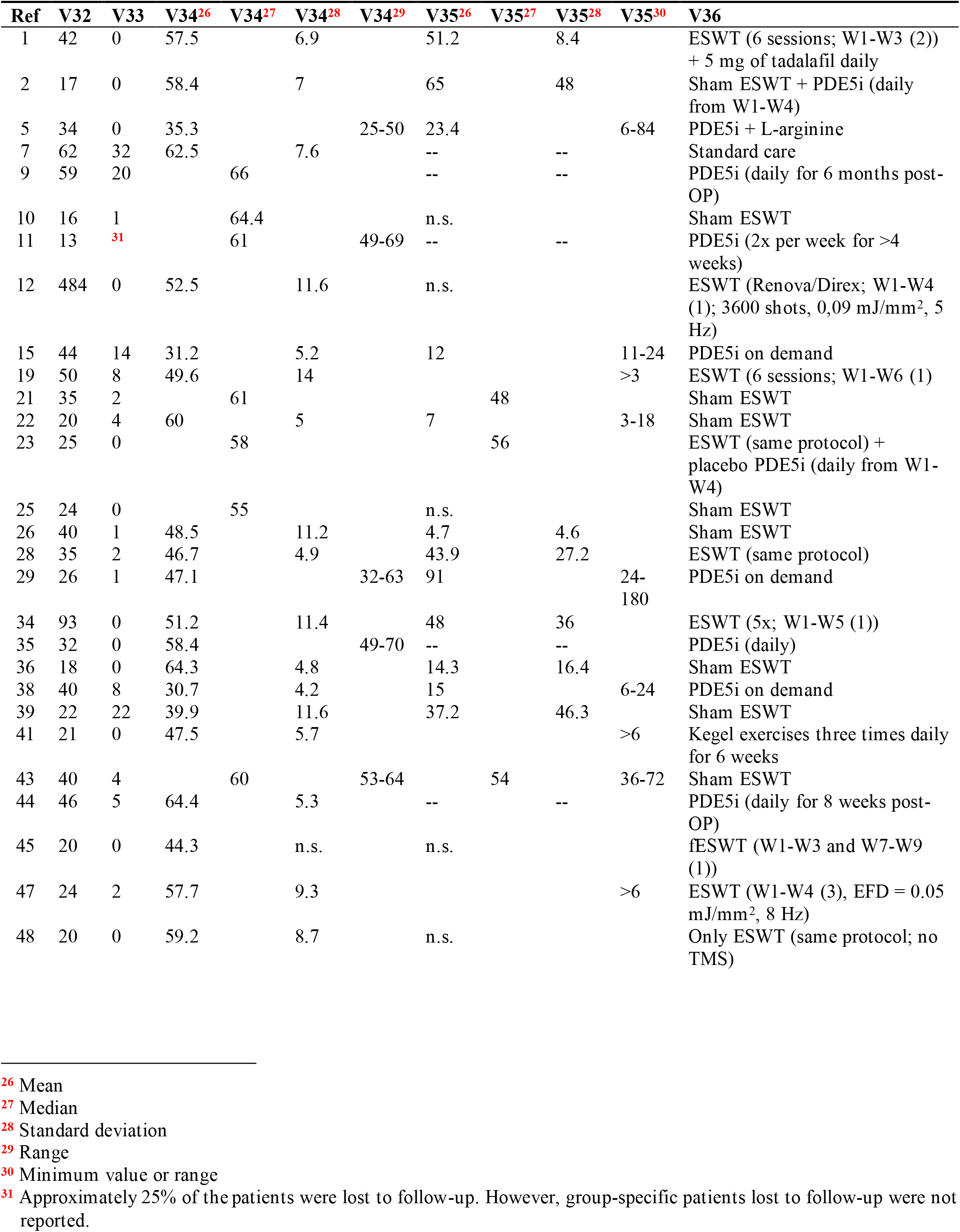
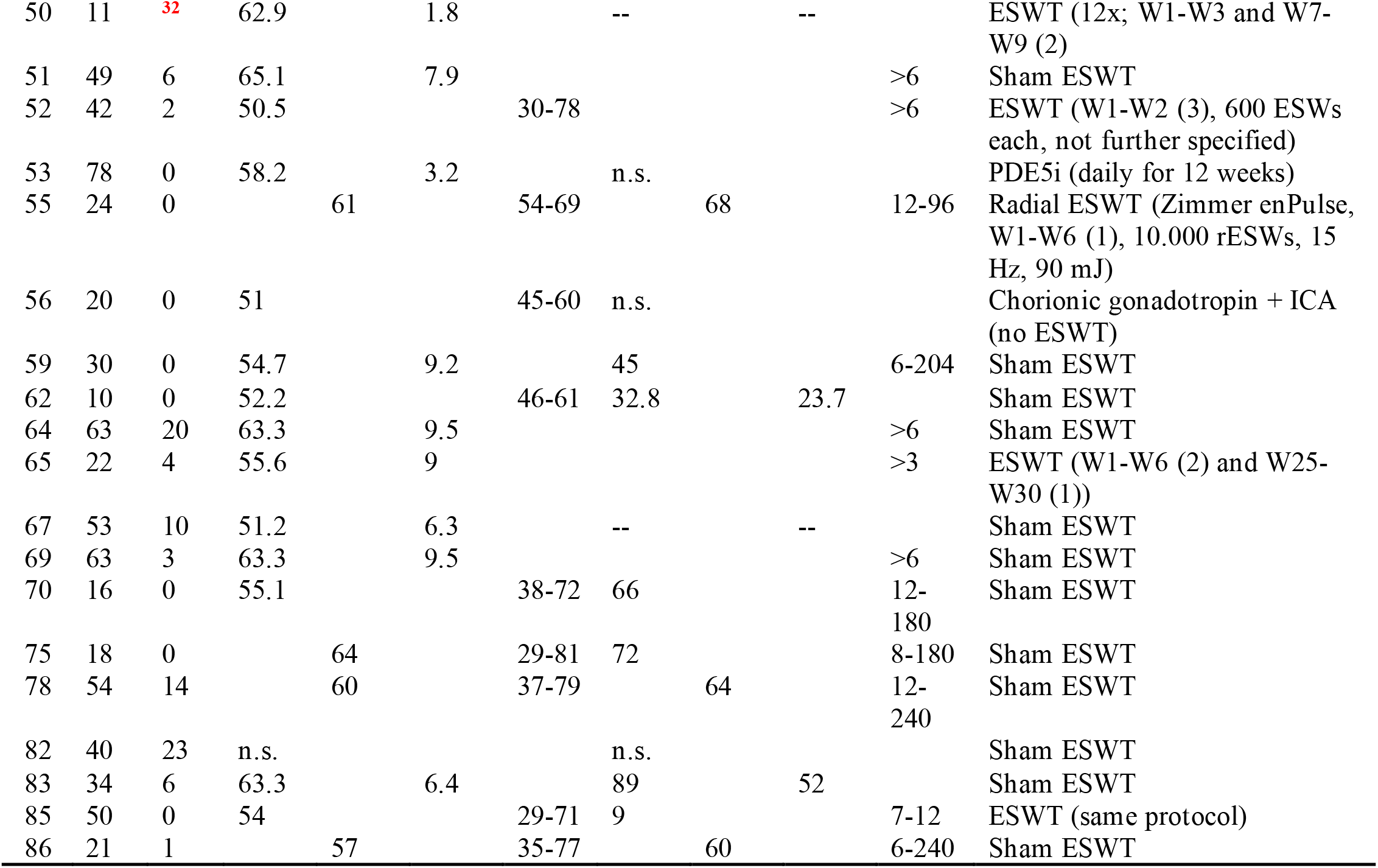

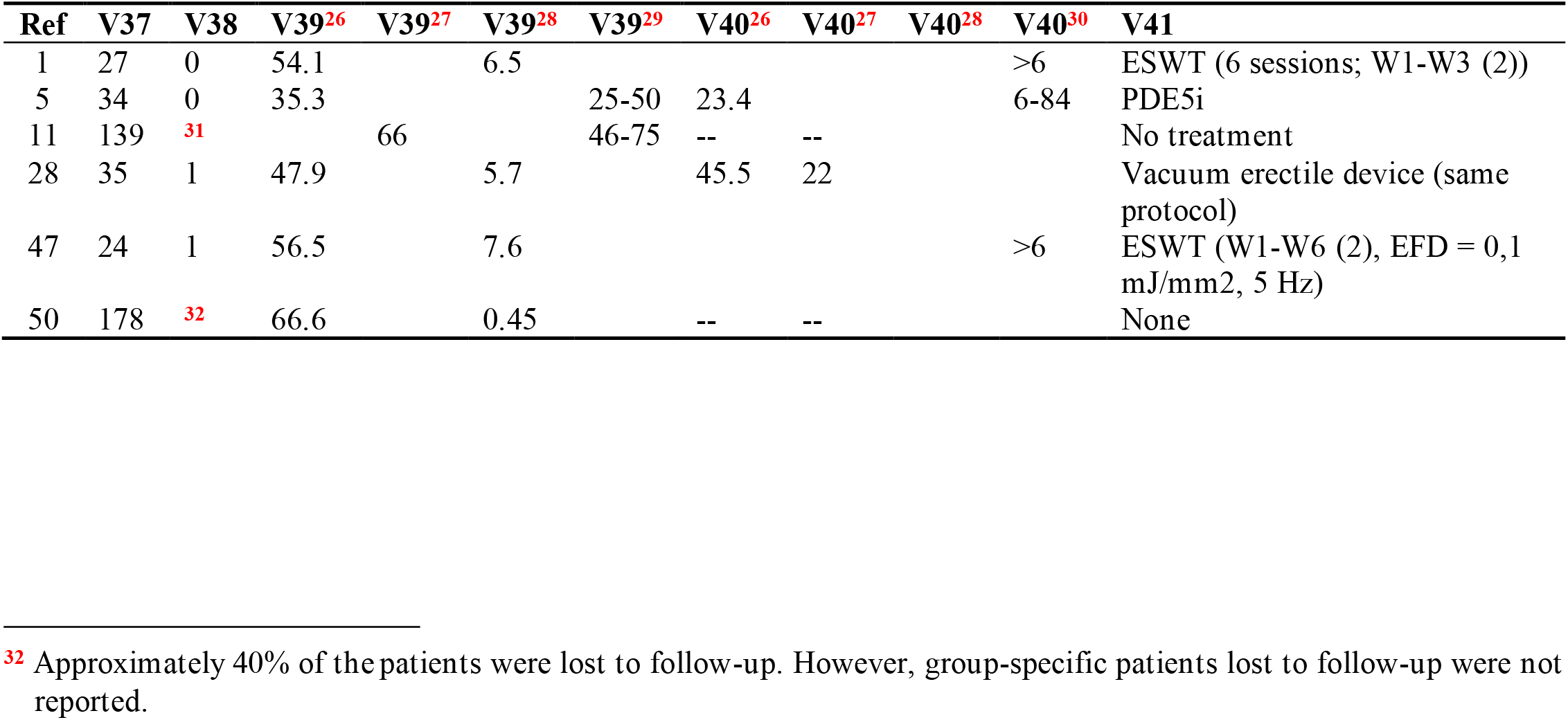
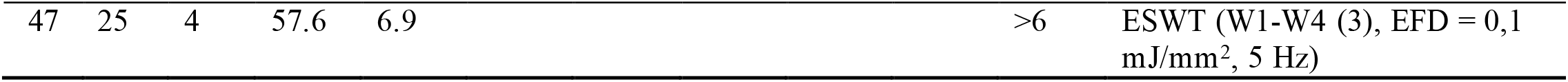

| Ref | V47 | V48 | V49 | V50 | V51 | V52<sup>33</sup> | V52<sup>34</sup> | V52<sup>35</sup> | V52<sup>36</sup> | V53 | V54 | V55 |
| --- | --- | --- | --- | --- | --- | --- | --- | --- | --- | --- | --- | --- |
| 1 | Yes | No | n.a. | n.a. | No |  |  |  |  | No | Yes |  |
| 2 | Yes | No | n.a. | n.a. | No | 7.7 | 1.3 | 12.9 | 2.1 | Yes | Yes | A |
| 3 | Yes | No | n.a. | n.a. | No |  |  |  |  | n.a. | Yes |  |
| 4 | Yes | No | n.a. | n.a. | No |  |  |  |  | n.a. | Yes |  |
| 5 | Yes | No | n.a. | n.a. | No | 14.7 | 1.5 | 25.8 | 3.6 | Yes | Yes | B |
| 6 | Yes | No | n.a. | n.a. | No |  |  |  |  | n.a. | Yes |  |
| 7 | No | Yes | No | n.a. | No |  |  |  |  | n.a. | n.a. |  |
| 8 | Yes | No | n.a. | n.a. | No |  |  |  |  | n.a. | Yes |  |
| 9 | No | Yes | No | n.a. | No |  |  |  |  | n.a. | n.a. |  |
| 10 | No | Yes | No | n.a. | No |  |  |  |  | Yes | Yes | A |
| 11 | No | Yes | No | n.a. | No |  |  |  |  | n.a. | n.a. |  |
| 12 | Yes | No | n.a. | n.a. | No |  |  |  |  | No | Yes |  |
| 13 | Yes | No | n.a. | n.a. | No |  |  |  |  | n.a. | Yes |  |
| 14 | No | Yes | No | n.a. | No | 14.3 | 5.2 | 20.3 | 8.4 | n.a. | Yes |  |
| 15 | No | Yes | No | n.a. | No | 8.6 | 3.6 | 16.3 | 5.5 | No | Yes | C |
| 16 | Yes | No | n.a. | n.a. | No |  |  |  |  | n.a. | Yes |  |
| 17 | Yes | No | n.a. | n.a. | No |  |  |  |  | n.a. | Yes |  |
| 18 | Yes | No | n.a. | n.a. | No |  |  |  |  | n.a. | No | B |
| 19 | No | Yes | No | n.a. | No | 16.5 | 4.1 | 21.5 | 4.5 | No | Yes |  |
| 20 | Yes | No | n.a. | n.a. | No | 15.17 | 3.75 | 21.61 | 3.6 | n.a. | Yes |  |
| 21 | No | Yes | No | n.a. | No | 14 | 1.7 | 20 | 2.4 | Yes | Yes | B |
| 22 | No | Yes | No | n.a. | No |  |  |  |  | Yes | n.a. | C |
| 23 | Yes | No | n.a. | n.a. | No | 21.2 | 2.5 | 25.3 | 2.3 | No | Yes |  |
| 24 | Yes | No | n.a. | n.a. | No |  |  |  |  | n.a. | Yes |  |
| 25 | Yes | No | n.a. | n.a. | No |  |  |  |  | Yes | Yes | A |
| 26 | Yes | Yes | Yes | BOCF | No | 16.2 | 0.6 | 17 | 1.2 | Yes | No | D |
| 27 | Yes | No | n.a. | n.a. | No |  |  |  |  | n.a. | Yes |  |
| 28 | No | Yes | No | n.a. | No | 13.3 | 1.6 | 18.9 | -- | No | Yes | A |
| 29 | No | Yes | No | n.a. | No |  |  |  |  | No | Yes | C |
| 30 | Yes | No | n.a. | n.a. | No |  |  |  |  | n.a. | Yes |  |
| 31 | No | Yes | No | n.a. | No |  |  |  |  | n.a. | Yes |  |
| 32 | Yes | No | n.a. | n.a. | No |  |  |  |  | n.a. | Yes |  |
| 33 | Yes | No | n.a. | n.a. | No |  |  |  |  | n.a. | No |  |
| 34 | Yes | No | n.a. | n.a. | No | 14.3 | 4.4 | 23.8 | 4.4 | No | Yes |  |
| 35 | Yes | No | n.a. | n.a. | No |  |  |  |  | Yes | n.a. | A |
| 36 | Yes | No | n.a. | n.a. | No |  |  |  |  | Yes | n.a. | A |
| 37 | No | Yes | No | n.a. | No |  |  |  |  | n.a. | Yes |  |
| 38 | No | Yes | No | n.a. | No |  |  |  |  | No | Yes | C |
| 39 | Yes | Yes | Yes | BOCF | No | 20.32 | 2.32 | 23.1 | 2.82 | No | Yes | A |
| 40 | No | Yes | No | n.a. | No | 13.5 | 4.6 | 22.1 | 5.3 | n.a. | Yes |  |
| 41 | Yes | No | n.a. | n.a. | No | 12.8 | 3.2 | 17.5 | 2.7 | Yes | Yes | A |
| 42 | Yes | No | n.a. | n.a. | No |  |  |  |  | n.a. | Yes |  |
| 43 | No | Yes | No | n.a. | No | 12 | 8-17<sup>37</sup> | 15 | 7-22<sup>37</sup> | No | Yes | A |
| 44 | No | Yes | No | n.a. | No |  |  |  |  | n.a. | n.a. |  |
| 45 | Yes | No | n.a. | n.a. | No |  |  |  |  | No | Yes |  |
| 46 | Yes | No | n.a. | n.a. | No |  |  |  |  | n.a. | Yes |  |
| 47 | No | Yes | No | n.a. | No | 18.3 | 4.2 | 23.6 | 4 | No | Yes |  |

<sup>33</sup> Mean LLEF-EF score at baseline
<sup>34</sup> Standard deviation of the LLEF-EF score at baseline
<sup>35</sup> Mean LLEF-EF score at follow-up
<sup>36</sup> Standard deviation of the LLEF-EF score at follow-up
<sup>37</sup> Range

| Ref | V47 | V48 | V49 | V50 | V51 | V52<sup>33</sup> | V52<sup>34</sup> | V52<sup>35</sup> | V52<sup>36</sup> | V53 | V54 | V55 |
| --- | --- | --- | --- | --- | --- | --- | --- | --- | --- | --- | --- | --- |
| 48 | Yes | No | No | n.a. | No |  |  |  |  | No | Yes |  |
| 49 | No | Yes | No | n.a. | No |  |  |  |  | n.a. | Yes |  |
| 50 | No | Yes | No | n.a. | No |  |  |  |  | n.a. | n.a. |  |
| 51 | No | Yes | No | n.a. | No | 16.6 | 3 | 21.7 | 3 | Yes | Yes | A |
| 52 | No | Yes | No | n.a. | No | 18.1 | 17.1-19.1<sup>38</sup> | 21 | 19.4-22.6<sup>38</sup> | No | Yes |  |
| 53 | Yes | No | n.a. | n.a. | No |  |  |  |  | Yes | Yes | B |
| 54 | Yes | No | n.a. | n.a. | No |  |  |  |  | n.a. | Yes |  |
| 55 | Yes | No | n.a. | n.a. | No |  |  |  |  | No | Yes |  |
| 56 | Yes | No | n.a. | n.a. | No |  |  |  |  | No | Yes |  |
| 57 | Yes | No | n.a. | n.a. | No |  |  |  |  | n.a. | Yes |  |
| 58 | No | Yes | No | n.a. | No |  |  |  |  | n.a. | Yes |  |
| 59 | Yes | No | n.a. | n.a. | No |  |  |  |  | Yes | Yes | A |
| 60 | Yes | No | n.a. | n.a. | No |  |  |  |  | n.a. | Yes |  |
| 61 | Yes | No | n.a. | n.a. | No |  |  |  |  | n.a. | Yes |  |
| 62 | Yes | No | n.a. | n.a. | No |  |  |  |  | Yes | Yes | A |
| 63 | Yes | No | n.a. | n.a. | No |  |  |  |  | n.a. | Yes |  |
| 64 | No<sup>39</sup> | Yes | No | n.a. | No | 11.2 | 6.6 | 16 | 9.8 | No | Yes |  |
| 65 | No | Yes | No | n.a. | No | 16.2 | 3.8 | 19.3 | 4 | No | Yes |  |
| 66 | Yes | No | n.a. | n.a. | No |  |  |  |  | n.a. | Yes |  |
| 67 | No | Yes | No | n.a. | No |  |  |  |  | n.a. | n.a. |  |
| 68 | No | Yes | No | n.a. | No |  |  |  |  | n.a. | Yes |  |
| 69 | No | Yes | No | n.a. | No | 10.9 | 9.1-12.7<sup>38</sup> | 13.1 | 9.3-13.4<sup>38</sup> | Yes | No | D |
| 70 | Yes | No | n.a. | n.a. | No | 13.8 | 3.6 | 19 | 3.3 | Yes | Yes | A |
| 71 | Yes | No | n.a. | n.a. | No |  |  |  |  | n.a. | Yes |  |
| 72 | No | Yes | No | n.a. | No |  |  |  |  | n.a. | Yes |  |
| 73 | No | Yes | No | n.a. | Yes<sup>40</sup> |  |  |  |  | n.a. | n.a. |  |
| 74 | Yes | No | n.a. | n.a. | No |  |  |  |  | n.a. | Yes |  |
| 75 | No | Yes | No | n.a. | No | 7 | 6-10<sup>37</sup> | 13 | 9-18<sup>37</sup> | No | Yes | A |
| 76 | Yes | No | n.a. | n.a. | No | 14.9 | -- | 19.9 | -- | n.a. | Yes |  |
| 77 | Yes | No | n.a. | n.a. | No |  |  |  |  | n.a. | No |  |
| 78 | No | Yes | No | n.a. | No |  |  |  |  | No | No |  |
| 79 | Yes | No | n.a. | n.a. | No | 15 | -- | 20 | -- | n.a. | Yes |  |
| 80 | Yes | No | n.a. | n.a. | No | 14.8 | 4.8 | 22.3 | -- | n.a. | Yes |  |
| 81 | Yes | No | n.a. | n.a. | No | 16.5 | 6.4 | 21 | 6.4 | n.a. | Yes |  |
| 82 | No | Yes | No | n.a. | No | 9.5 | 2 | 22 | 3 | Yes | Yes | A |
| 83 | No | Yes | No | n.a. | No | 10.2 | 3.8 | 17.8 | 4.8 | Yes | Yes | C |
| 84 | No | Yes | No | n.a. | No | 8.8 | -- | 12.3 | -- | n.a. | Yes |  |
| 85 | Yes | No | n.a. | n.a. | No | 11.6 |  | 21.5 |  | No | Yes | B |
| 86 | No | Yes | No | n.a. | No | 11.5 | 0.86 | 18.2 | 0.8 | Yes | Yes | B |
| 87 | Yes | No | n.a. | n.a. | No | 13.5 | 4.1 | 20.9 | 5.8 | n.a. | Yes |  |

<sup>38</sup> 95% confidence interval of the mean
<sup>39</sup> The authors stated that they performed a modified ITT analysis. To this end, patients who were found ineligible after randomization were excluded (i.e. patients who met all inclusion criteria but with a baseline IIEF-EF score > 25). However, an IIEF-EF score $\geq 25$ was an exclusion criteria used in this trial, raising the question why patients were randomized into the trial despite the fact that they fulfilled one of the exclusion criteria and were therefore excluded after randomization. Furthermore, patients with missing primary outcome data were also excluded. However, this should be considered lack of missing data imputation rather than performing a modified ITT analysis.
<sup>40</sup> Patients with de novo use of erectogenic aids at the first follow-up were excluded from the analysis.

